# Interpretable machine learning prediction of in-hospital mortality in ICU patients with cancer and sepsis using first-day data: Development using MIMIC-IV and external validation in eICU-CRD

**DOI:** 10.64898/2026.07.30.26359086

**Authors:** Janet Sanjaya, Sakshie Pathak, Yong Si, Mohammadsaeed Haghi, Nausin Kudrot, Greg Placencia, Kamiar Alaei, Maryam Pishgar

**Affiliations:** Daniel J. Epstein Department of Industrial and Systems Engineering, University of Southern California, Los Angeles, 90089, California, USA; Department of Electrical and Electronic Engineering, Bangladesh University of Engineering and Technology, Dhaka, Bangladesh; Industrial and Manufacturing Engineering, California State Polytechnic University, Pomona, Pomona, 91768, California, USA; Department of Health Science, California State University, Long Beach, Long Beach, 90840, California, USA

**Keywords:** Cancer, Sepsis, Intensive care unit, Machine learning, Mortality prediction, External validation, Explainable artificial intelligence

## Abstract

**Background:** Critically ill patients with cancer and sepsis have high in-hospital mortality, but externally validated prediction models are limited.

**Objective:** To develop and externally validate an interpretable machine learning framework using first-day intensive care data.

**Methods:** We used MIMIC-IV version 3.1 for development and internal validation and eICU-CRD for external validation. Eligible adults had cancer, an intensive care unit stay of at least 24 hours, and met a prespecified operational sepsis definition. The prediction landmark was 24 hours after admission. The MIMIC-IV cohort included 3,729 stays (training, *n* = 2, 983; internal validation, *n* = 746). Same-admission diagnosis-derived variables were excluded, and 345 predictors were retained. Fourteen predictive models and a dummy baseline were evaluated. Frozen pipelines and training-derived thresholds were applied to eICU-CRD without refitting or recalibration.

**Results:** Gradient boosting was selected as the primary model and achieved an internal AUROC of 0.8480 (95% CI, 0.8173–0.8755), AUPRC of 0.6984, and Brier score of 0.1346. Important predictors included Glasgow Coma Scale components, temperature, lactate dehydrogenase, age, respiratory rate, oxygen saturation, blood urea nitrogen, and serum lactate. In eICU-CRD (*n* = 611), gradient boosting achieved an AUROC of 0.7483 (95% CI, 0.7026–0.7919), AUPRC of 0.6243, and Brier score of 0.1709. Random forest had the highest external AUROC in secondary comparisons (0.7731).

**Conclusions:** First-day data supported useful internal discrimination, but performance declined under locked external validation. Multicenter validation, recalibration, threshold assessment, and prospective evaluation are required before clinical implementation.

## 1. Introduction

Cancer remains a major cause of morbidity and mortality worldwide. In 2020, an estimated 19.3 million new cancer cases and nearly 10.0 million cancer deaths occurred globally, and approximately one in five people develop cancer during their lifetime [1]. As cancer incidence increases and survival after diagnosis improves, a growing number of patients with malignancy require intensive care for acute complications of the disease or its treatment. Sepsis, defined as life-threatening organ dysfunction caused by a dysregulated host response to infection, is a particularly important complication in this population [2]. Cancer-related immunosuppression, cytotoxic treatment, mucosal barrier injury, invasive devices, and reduced physiological reserve increase susceptibility to severe infection. Severe sepsis has been associated with a substantial proportion of deaths among hospitalized patients with cancer [3]. Systematic reviews and multicenter studies have also demonstrated persistently high short-term mortality among critically ill patients with cancer and sepsis, although outcomes vary substantially according to study period, malignancy characteristics, organ dysfunction, functional status, and the requirement for life-sustaining treatment [4, 5, 6]. These findings underscore the need for reliable and population-specific risk stratification.

The clinically relevant task is not limited to identifying sepsis, but also includes estimating the risk of subsequent in-hospital death among patients who have both cancer and sepsis and remain critically ill after initial stabilization. Conventional severity scores, including the Acute Physiology and Chronic Health Evaluation, Simplified Acute Physiology Score, and Sequential Organ Failure Assessment, provide useful summaries of physiological derangement and organ dysfunction. However, their performance varies across critically ill oncology populations, and they may not fully represent cancer-specific prognostic factors such as malignancy type, metastatic disease, treatment-related immunosuppression, hematologic disease, and altered physiological reserve [7, 8]. A prediction framework developed specifically for patients with cancer and sepsis may therefore provide more relevant risk estimates and may help identify patients requiring closer monitoring and reassessment.

Previous studies have evaluated mortality prediction in sepsis and in critically ill patients with cancer. Yuan et al. developed a LASSO-selected logistic regression nomogram for hospital mortality among critically ill patients with sepsis and solid cancer using MIMIC-IV and externally validated the model using eICU-CRD, reporting areas under the receiver operating characteristic curve of 0.726 and 0.756 in the internal and external validation cohorts, respectively [9]. More recently, Tang et al. evaluated interpretable machine learning algorithms in patients with lung cancer and sepsis using a population-based development dataset and an external validation cohort [10]. These studies support the feasibility of population-specific mortality prediction but remain focused on selected malignancy subgroups.

In broader sepsis populations, Hou et al. reported that XGBoost outperformed logistic regression and SAPS II for 30-day mortality prediction using MIMIC-III [11]. Li et al. developed an interpretable machine learning model for in-hospital mortality using MIMIC-IV and identified age, cancer, and cardiovascular comorbidities among influential predictors [12]. Wang et al. similarly compared multiple algorithms for early sepsis mortality prediction and reported favorable performance for an interpretable machine learning framework [13]. Bao et al. developed sepsis mortality models using MIMIC-IV and evaluated their performance in eICU-CRD, providing a relevant cross-database benchmark [14]. Multicenter sepsis studies have also incorporated clinical and inflammatory biomarkers to evaluate model performance across heterogeneous patient populations [15].

Despite these advances, several gaps remain. First, studies specifically addressing patients with concurrent cancer and sepsis are limited, and the most directly comparable externally validated studies have focused primarily on solid cancer or a single malignancy type [9, 10]. Second, evidence that machine learning models consistently outperform well-specified regression models in clinical prediction remains mixed [16]. Comparisons across model families are difficult to interpret when preprocessing, feature selection, tuning, and evaluation procedures differ between algorithms. Third, prediction studies frequently emphasize discrimination while providing less complete assessment of precision–recall performance, calibration, probabilistic accuracy, and clinically relevant operating thresholds. These complementary dimensions are important because a model with favorable discrimination may still produce poorly calibrated probabilities or unsuitable classifications at a selected threshold [17, 18]. Finally, internal performance may not be preserved when a model is transferred to populations with different case mix, coding practices, measurement patterns, predictor availability, and care processes [19, 20, 21].

In this study, we developed and externally evaluated an interpretable machine learning framework for predicting subsequent in-hospital mortality among adult intensive care unit patients with cancer and sepsis. Model development and internal validation were performed using MIMIC-IV version 3.1, and external validation was performed using eICU-CRD [22, 23]. Because time-varying predictors were summarized over the first 24 hours after intensive care unit admission, the prediction landmark was defined as 24 hours after admission. The resulting models therefore estimate subsequent in-hospital mortality among patients who remained in intensive care through this landmark rather than mortality risk at the moment of admission.

The study included both solid and hematologic malignancies and integrated demographic, admission, vital-sign, Glasgow Coma Scale, and laboratory variables. Same-admission diagnosis-derived variables were excluded from modeling because their availability by the 24-hour prediction landmark could not be established. Fourteen predictive algorithms and a dummy baseline were evaluated under shared preprocessing and cross-validated hyperparameter tuning. Model performance was assessed using discrimination, precision– recall performance, threshold-dependent classification metrics, probabilistic prediction error, and graphical calibration. SHapley Additive exPlanations were used to examine the internally selected model [24]. External validation was conducted by applying the frozen MIMIC-IV preprocessing pipelines, model parameters, feature set, and operating thresholds to eICU-CRD without model refitting, feature reselection, or recalibration. Study reporting was guided by the TRIPOD+AI statement, while potential risks of bias and applicability concerns were considered with reference to PROBAST+AI [25, 26].

## 2. Materials and methods

### 2.1. Data sources and ethics

This retrospective study used the Medical Information Mart for Intensive Care IV database (MIMIC-IV, version 3.1) for model development and internal validation [22]. MIMIC-IV contains de-identified health-related data from patients admitted to intensive care units at Beth Israel Deaconess Medical Center in Boston, Massachusetts, United States, between 2008 and 2019. Available data include patient demographics, hospital admissions, diagnosis codes, laboratory measurements, microbiology records, medication information, vital signs, and derived clinical variables.

The eICU Collaborative Research Database (eICU-CRD, version 2.0) was used exclusively for external validation [23]. eICU-CRD is a multicenter critical care database containing de-identified records from more than 200,000 intensive care admissions across 208 hospitals in the United States during 2014 and 2015.

Data were extracted using structured query language in Google BigQuery. Cohort construction and first-day feature generation followed a three-layer pipeline comprising: (1) construction of the base cohort and relevant static variables; (2) generation of first-24-hour laboratory and physiological summaries; and (3) assembly of the patient–intensive-care-stay-level modeling datasets. The resulting tables were exported and processed in Python.

Access to MIMIC-IV and eICU-CRD was obtained through PhysioNet after completion of the required credentialing and human-subjects research training. Both databases contain de-identified data collected under their respective institutional approvals and data-use agreements. Because this study involved secondary analysis of de-identified records and no direct interaction with human participants, no additional institutional review board approval or individual informed consent was required.

### 2.2. Study population and cohort construction

Adult intensive care unit patients with cancer and sepsis were retrospectively identified in MIMIC-IV version 3.1 using prespecified eligibility criteria. Patients were required to have a MIMIC-IV anchor age of at least 18 years and to have an intensive care unit length of stay of at least 24 hours. Because time-varying predictors were summarized during the first 24 hours after intensive care unit admission, the prediction landmark was defined as 24 hours after admission. The resulting study population therefore comprised patients who remained in intensive care through the landmark, and the models should not be interpreted as admission-time prediction models.

Cancer status was identified using International Classification of Diseases, Ninth Revision (ICD-9), and Tenth Revision (ICD-10), diagnosis codes associated with the index hospitalization. Solid malignancy was defined using ICD-9 codes 140–199 or 209 and ICD-10 codes C00–C80, C7A, or C7B. Hematologic malignancy was defined using ICD-9 codes 200–208 and ICD-10 codes C81–C96. These diagnosis codes were used retrospectively to define cohort eligibility. For each patient, the earliest cancer-associated intensive care stay meeting the age and length-of-stay requirements was retained and subsequently assessed using the operational sepsis definition.

Sepsis was identified using a prespecified operational definition based on evidence of suspected infection around intensive care admission and laboratory evidence of organ dysfunction during the first 24 hours. This database-compatible definition was used to support comparable cohort construction in MIMIC-IV and eICU-CRD and was not intended to represent a complete implementation of the Sequential Organ Failure Assessment-based Sepsis-3 criteria [2].

For stay *i*, the infection-ascertainment window was defined as

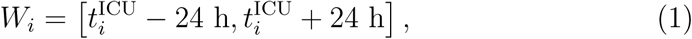

where 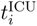 denotes the intensive care admission time. Evidence of suspected infection was coded as

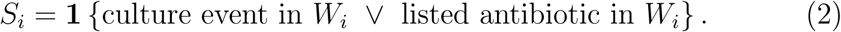

Here “listed antibiotic” denotes a medication matching the prespecified antibiotic name list. Laboratory evidence of organ dysfunction during the first 24 hours was coded as

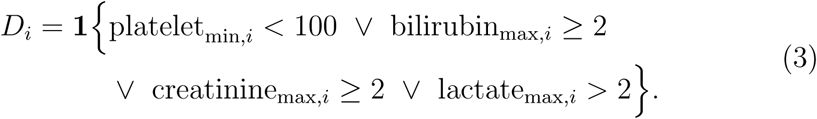

where platelet count was measured in 10^9^*/*L, bilirubin and creatinine in mg*/*dL, and serum lactate in mmol*/*L. In MIMIC-IV, the lactate criterion was restricted to serum lactate item 50813; lactate dehydrogenase was not used for cohort construction. In eICU-CRD, the corresponding criterion used the exact laboratory concept “lactate.” The operational sepsis indicator was then defined as

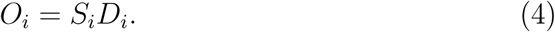

Stays with *O_i_* = 1 were retained. The MIMIC-IV cohort comprised 3,729 unique intensive care stays, including 1,027 deaths during the index hospitalization and 2,702 survivors to hospital discharge. The cohort was divided using an 80:20 stratified random split into a training set (*n* = 2, 983; 822 deaths) and an internal validation set (*n* = 746; 205 deaths), preserving hospital mortality prevalence across the two partitions.

Cohort construction is summarized in Figure 1.

**Figure 1:**
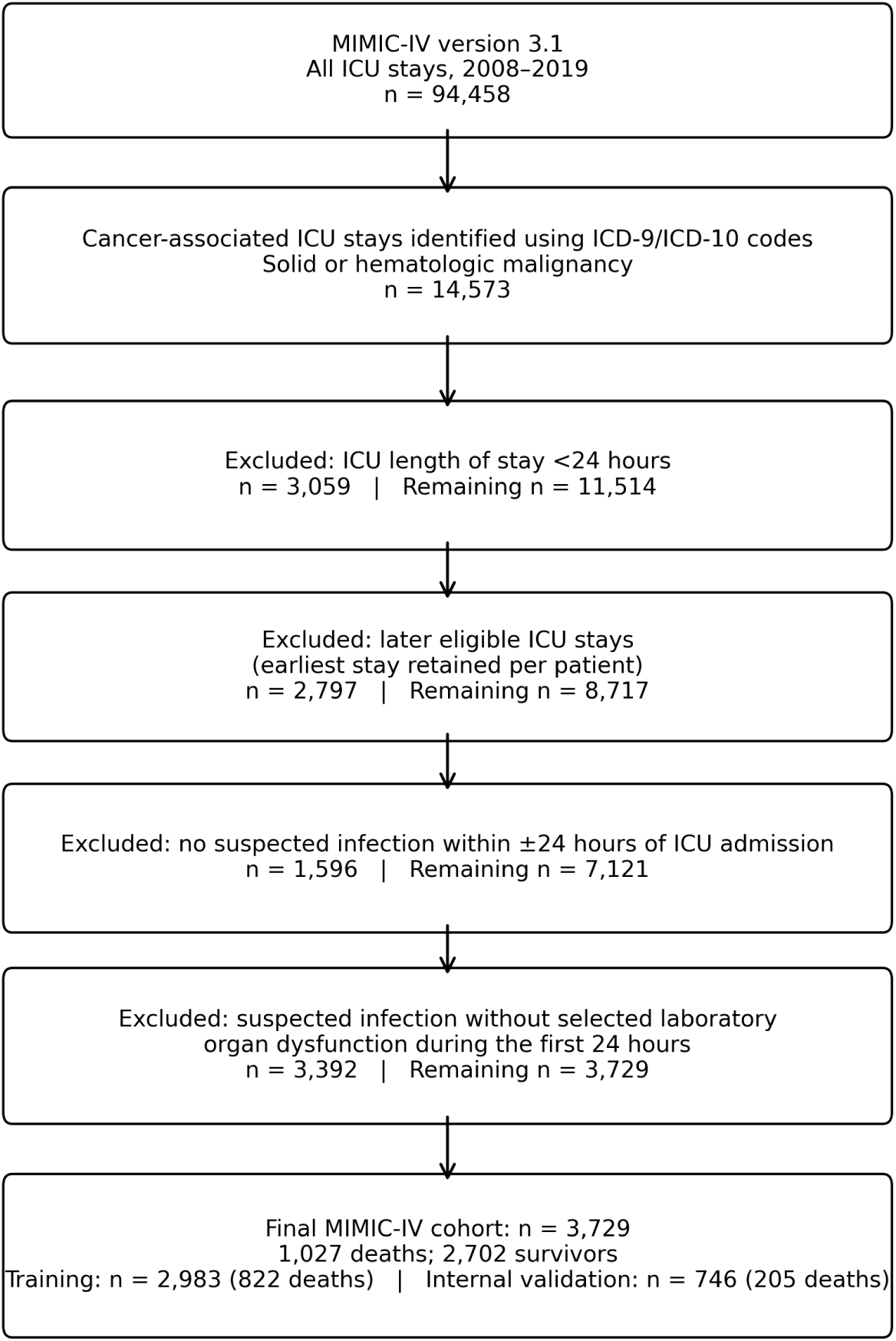
Cohort construction for adult intensive care unit patients with cancer and sepsis in MIMIC-IV version 3.1. For each patient, the earliest cancer-associated intensive care stay meeting the age and length-of-stay requirements was retained and subsequently assessed using the prespecified operational sepsis definition. Eligible stays were then divided into stratified training and internal validation sets.

### 2.3. Outcome definition

The primary outcome was subsequent in-hospital mortality after the 24-hour prediction landmark during the index hospitalization. Outcome status was obtained from the MIMIC-IV `hospital_expire_flag` field and was coded as

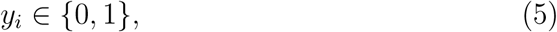

where *y_i_* = 1 indicated death before hospital discharge and *y_i_* = 0 indicated survival to hospital discharge. Records without a valid outcome value were excluded before the development cohort was divided into training and internal validation sets.

### 2.4. Predictor variables

Candidate predictors were derived from structured MIMIC-IV records and were grouped into demographic and admission characteristics, vital signs, Glasgow Coma Scale components, and laboratory-related variables. Time-varying physiological and laboratory predictors were restricted to measurements recorded during the first 24 hours after intensive care unit admission. Static demographic variables included age, sex, race, marital status, insurance, language, admission type, admission location, and first care unit.

Same-admission diagnosis-derived variables were excluded from the modeling matrices because MIMIC-IV hospital diagnosis codes are generally assigned at the hospitalization level and the eICU-CRD diagnosis extraction included records from the complete unit stay. Their availability by the 24-hour prediction landmark could therefore not be established consistently. The excluded variables comprised solid- and hematologic-malignancy indicators, 14 diagnosis-derived comorbidity indicators, the number of comorbidity flags, a Charlson-like score, the number of index-hospitalization diagnosis records, and the number of unique index-hospitalization diagnosis codes. Cancer diagnosis codes were retained solely for retrospective cohort eligibility.

Vital-sign predictors included heart rate, systolic blood pressure, diastolic blood pressure, mean blood pressure, respiratory rate, peripheral oxygen saturation, and temperature. Each vital sign was summarized over the first 24 hours using the minimum, maximum, and mean values.

Glasgow Coma Scale eye, verbal, and motor components were summarized using the minimum, maximum, mean, first, last, and measurement-count statistics. Laboratory predictors were generated from raw laboratory records obtained during the first 24 hours. For laboratory item *ℓ* and stay *i*, the summary vector was

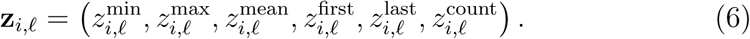

No laboratory ratios, temporal slopes, or explicit interaction terms were constructed beyond these summary features. Details of train-only feature filtering and the final predictor composition are provided below.

### 2.5. Data preprocessing

The development cohort was first divided into stratified training and internal validation sets. Identifiers, direct outcome-related variables, operational cohort-definition indicators, and the prespecified diagnosis-derived predictors were removed. Train-only missingness and variance filtering was then applied before plausibility cleaning. Values outside prespecified physiologically credible ranges were subsequently set to missing, categorical variables were recoded, and the grouped admission-type indicator was added. Because plausibility cleaning occurred after the missingness filter, values converted to missing by the plausibility rules did not contribute to the greater-than-70% missingness exclusions.

Patient, hospital-admission, and intensive-care-stay identifiers were excluded before modeling. Variables considered to introduce direct outcome leakage or tautology were also removed, including death and discharge timestamps, length-of-stay variables, last care unit, and the operational cohort-definition indicators for suspected infection, organ dysfunction, and sepsis.

Sex was recoded as Female, Male, or Other. Race categories were collapsed into Asian, Black, White, and Other. Admission type was additionally grouped as Emergency or Elective. Identical recoding rules were used in the training and internal validation sets.

Prespecified plausibility rules were used to identify values considered incompatible with physiologically credible measurements or likely to represent recording errors. Values outside the following ranges were set to missing: heart rate, 10–300 beats/min; systolic blood pressure, 30–350 mmHg; diastolic blood pressure, 10–250 mmHg; mean blood pressure, 10–300 mmHg; respiratory rate, 1–80 breaths/min; peripheral oxygen saturation, 50–100%; and temperature, 30–45°C. Glasgow Coma Scale eye, verbal, and motor values outside their valid ranges of 1–4, 1–5, and 1–6, respectively, were also set to missing. Age outside 18–120 years, glucose above 1,000 mg/dL, lactate values less than or equal to 0 or above 30 mmol/L, and clearly invalid coagulation measurements were handled in the same manner. Pathologically extreme but clinically possible observations were retained, and no post-imputation winsorization or clipping was performed.

Predictors were classified as numeric or categorical. Missing numeric values were imputed using the median estimated from the training data. Numeric predictors were then standardized as

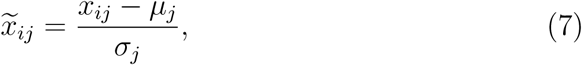

where *µ_j_* and *σ_j_* were estimated using the training set and applied unchanged to the internal validation set. Missing categorical values were assigned to an “Unknown” category, after which categorical variables were one-hot encoded. Unseen categories in the validation set were ignored during transformation.

Imputation, categorical encoding, and numeric scaling were implemented using a `ColumnTransformer` within each model pipeline. This design ensured that all preprocessing parameters were learned exclusively from the corresponding training data during cross-validation and final model fitting. Restricting preprocessing estimation to the training data was intended to reduce information leakage and optimistic performance estimation. No over-sampling or under-sampling procedure was used. Class-weighted objectives were used for selected classifiers that supported this option.

### 2.6. Feature filtering

Feature filtering was performed exclusively using the training partition and did not use outcome labels. For candidate predictor *j*, the training-set missingness proportion was defined as

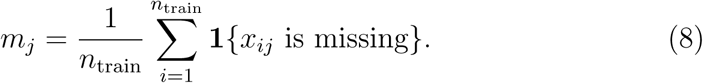

Predictors with more than 70% missingness in the training set were excluded:

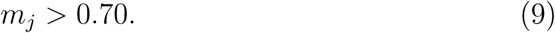

All-missing variables and variables with no variation in the training set were also removed. The set of retained variables was then applied unchanged to the internal validation data. No univariate outcome-based screening, recursive feature elimination, or embedded supervised feature-selection procedure was used.

The initial matrix contained 2,702 candidate predictors after removal of identifiers, leakage-related fields, and diagnosis-derived variables. Train-only missingness filtering removed 2,358 predictors. No additional predictors were removed by the all-missing or zero-variance filters, leaving 344 retained predictors. Addition of the grouped Emergency-versus-Elective admission indicator produced the final set of 345 predictors. This same fixed feature set was used for model development, internal evaluation, interpretation, and external scoring.

### 2.7. Machine learning models

Fourteen predictive classifiers and one non-informative dummy baseline were evaluated using the same development cohort, feature set, and preprocessing framework. The evaluated methods were:

- most-frequent dummy classifier;
- ridge logistic regression;
- class-balanced ridge logistic regression;
- LASSO logistic regression;
- class-balanced LASSO logistic regression;
- elastic-net logistic regression;
- decision tree;
- random forest;
- extremely randomized trees;
- gradient boosting;
- histogram-based gradient boosting;
- AdaBoost;
- Gaussian naive Bayes;
- *k*-nearest neighbors; and
- multilayer perceptron.

Logistic regression models served as linear baselines. Decision trees, bagged tree ensembles, and boosting models were included to capture nonlinear predictor relationships and interactions. Gaussian naive Bayes, *k*-nearest neighbors, and the multilayer perceptron provided additional classical and neural-network comparators.

For logistic regression, the predicted probability of in-hospital death was expressed as

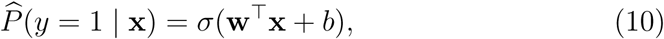

where

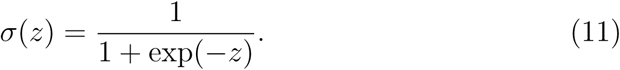

Regularization penalties differed across the ridge, LASSO, and elastic-net variants. Models supporting class weighting used balanced class weights where specified, including the balanced logistic regression variants and selected tree-based classifiers. Each estimator was fitted within a pipeline containing the shared preprocessing transformer and the corresponding classifier. Model pipelines, preprocessing transformers, and grid-search procedures were implemented using scikit-learn [27].

### 2.8. Hyperparameter tuning

Hyperparameters were selected using grid search with five-fold stratified cross-validation within the training set. Cross-validation folds were generated using

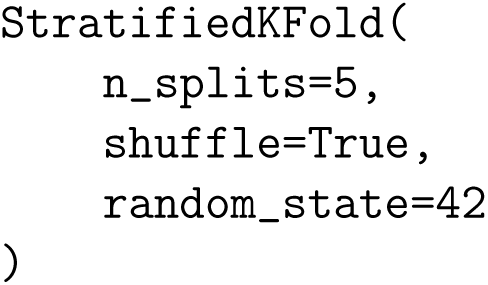

and the mean cross-validated area under the receiver operating characteristic curve was used as the optimization criterion. For model family *M* with hyperparameter configuration ***θ***, the tuning objective was

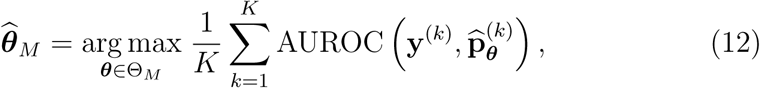

where *K* = 5, Θ*_M_* denotes the prespecified parameter grid, and denotes 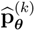 predictions for the held-out fold *k*.

The search spaces included regularization strength and penalty parameters for logistic regression, tree depth and minimum leaf size for tree-based models, numbers of estimators and learning rates for boosting methods, neighborhood size for *k*-nearest neighbors, and hidden-layer configurations for the multilayer perceptron. After selection of the optimal configuration, `GridSearchCV` refitted each model using the complete training set. Random seeds were fixed to support reproducibility.

### 2.9. Operating-threshold selection

Threshold-independent model selection was based primarily on internal-validation AUROC. Threshold-dependent classification metrics were calculated using a model-specific operating threshold selected exclusively from cross-validated training predictions.

For each model, out-of-fold predicted probabilities were generated within the training set. The operating threshold was selected by maximizing Youden’s index:

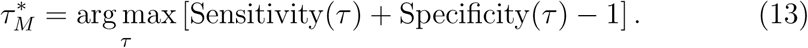

When multiple thresholds produced the same maximum value, the first maximizing threshold was retained. The resulting threshold was applied unchanged to the held-out internal validation set and, subsequently, to the external validation cohort. Neither the internal validation outcomes nor the external outcomes were used to re-estimate the operating threshold.

### 2.10. Model evaluation and statistical analysis

Model performance was evaluated primarily on the held-out internal validation set. Performance was assessed across complementary domains rather than using discrimination alone, consistent with established recommendations for evaluating clinical prediction models [17]. Discrimination was quantified using the area under the receiver operating characteristic curve (AUROC) and the area under the precision–recall curve (AUPRC). AUROC was defined as

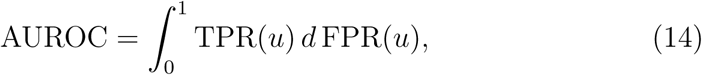

where TPR and FPR denote the true-positive and false-positive rates, respectively.

Threshold-dependent performance was summarized using accuracy, balanced accuracy, precision, sensitivity, specificity, and the F1 score. These metrics were calculated using the model-specific operating thresholds estimated from training-set out-of-fold predictions, as described above. The internal validation set was not used for threshold selection.

Probabilistic performance was assessed using the Brier score,

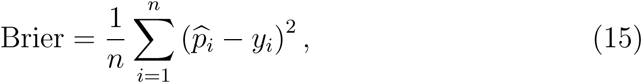

where 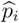 denotes the predicted probability of in-hospital death and *y_i_* denotes the observed outcome. Calibration was examined graphically using reliability curves constructed with 10 quantile-based bins. Lower Brier scores indicated lower overall prediction error, but were interpreted jointly with discrimination and graphical calibration rather than as a complete measure of calibration [18].

The primary model was selected according to the highest AUROC on the held-out internal validation set. AUPRC and Brier score were considered complementary performance measures. External cohort results were not used to select or retune the primary model.

Ninety-five percent confidence intervals for AUROC were estimated using ordinary nonparametric bootstrap resampling with 1,000 bootstrap samples drawn with replacement and a fixed random seed of 42. The 2.5th and 97.5th percentiles of the bootstrap AUROC distribution were reported. Resamples containing only one outcome class were discarded.

Baseline characteristics were summarized separately for the MIMIC-IV and eICU-CRD cohorts. Continuous variables were described using the median and interquartile range or the mean and standard deviation, as appropriate. Between-cohort comparisons used the Mann–Whitney *U* test for continuous variables and the chi-square test for categorical variables. Fisher’s exact test was used when expected cell counts were small. All tests were two-sided. Because these comparisons were descriptive and involved multiple variables, the resulting *p*-values were interpreted as indicators of between-cohort differences rather than confirmatory hypothesis tests.

### 2.11. Model interpretation

The internally selected primary model was interpreted using SHapley Additive exPlanations (SHAP) [24]. Because the selected model was tree-based, `TreeExplainer` was used to estimate feature-level contributions to individual predictions.

A stratified sample of 100 observations from the training set was used as the background dataset, and a stratified sample of 300 observations from the held-out internal validation set was used for explanation. For predictor *j*, global feature importance was calculated as the mean absolute SHAP value:

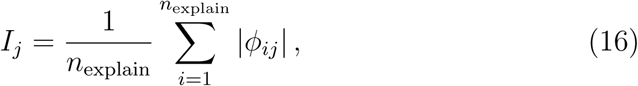

where *ϕ_ij_* denotes the SHAP contribution of predictor *j* for observation *i*, and *n*_explain_ = 300.

The 20 predictors with the highest mean absolute SHAP values were displayed using a global importance bar plot and a beeswarm summary plot. SHAP values were interpreted as model-specific contributions to predictions rather than causal effects, biological mechanisms, or evidence of clinical actionability [28]. The analysis was intended to examine whether the model relied on clinically interpretable patterns and to identify variables contributing most strongly to variation in predicted mortality risk.

### 2.12. External validation

External validation was performed using an independently constructed cohort from eICU-CRD. The external analysis was designed to assess model transportability rather than to provide an additional stage of model development [19, 20, 21]. Before external scoring, a locked development bundle was exported from the MIMIC-IV analysis. This bundle contained the final retained feature set, training-fitted preprocessing and model pipelines, selected hyperparameters, and model-specific operating thresholds estimated from MIMIC-IV training-set out-of-fold predictions.

The eICU-CRD cohort was constructed using criteria designed to approximate those applied in MIMIC-IV. Patients were required to be adults, to have an intensive care unit stay of at least 24 hours, and to meet the cancer and operational sepsis definitions. For each patient, the earliest cancer-associated intensive care stay meeting the age and length-of-stay requirements was retained and subsequently assessed using the operational sepsis definition. Because ages greater than 89 years are de-identified in eICU-CRD, recorded ages above 89 years were represented as 89 years in the external analysis.

Evidence of suspected infection was identified using microbiology culture records or receipt of a medication included in the prespecified antibiotic name list within 24 hours before or after unit admission. Laboratory evidence of organ dysfunction was assessed using measurements obtained during the first 24 hours after admission. The same demographic recoding and plausibility-screening rules used in the development cohort were applied to the external data.

In the final external pipeline, laboratory concepts were harmonized before locked scoring through the BigQuery extraction and subsequent preparation of the analysis matrix. Blood-gas pH was aligned to MIMIC-IV item 50820, and standard red-cell distribution width was aligned to item 51277. Ionized-calcium values greater than 2 were converted to the MIMIC-compatible mmol/L scale by division by 4, whereas values of 2 or less were retained without conversion, before alignment to free-calcium item 50808. No additional laboratory-name remapping was applied after construction of the aligned external analysis matrix. Glasgow Coma Scale eye, verbal, and motor components were extracted from first-day nurse-charting records; all three components were available for 452 of 611 external stays (74.0%).

External variables were aligned to the fixed 345-predictor development feature set. Unavailable features remained missing and were processed using imputation parameters learned from the MIMIC-IV training set. Thirty-nine of the 345 predictors were entirely missing in eICU-CRD. Mean feature-level missingness was 36.6% in eICU-CRD and 20.2% in the MIMIC-IV modeling data.

For each model, the frozen MIMIC-IV pipeline was applied directly to the aligned eICU-CRD matrix to generate mortality probabilities. No feature reselection, imputation refitting, encoder refitting, scaler refitting, hyperparameter tuning, model refitting, recalibration, or operating-threshold re-estimation was performed using eICU-CRD. External performance was evaluated using AUROC, AUPRC, Brier score, precision, sensitivity, specificity, F1 score, balanced accuracy, and accuracy.

GradientBoosting, selected according to internal validation AUROC before examination of the external results, remained the prespecified primary model for external reporting. Performance of the remaining models was reported as a secondary comparative analysis. A model that performed better on eICU-CRD was not retrospectively substituted as the primary model, because doing so would constitute model selection using the external validation cohort.

This locked evaluation design was intended to provide a direct assessment of performance under changes in case mix, documentation, measurement patterns, and predictor availability rather than an estimate optimized for the external cohort [19, 21].

### 2.13. Software

Data processing, model development, statistical evaluation, and visualization were performed in Python. Machine learning pipelines were implemented using scikit-learn [27], and model interpretation was performed using the SHAP package [24]. Random seeds were fixed where supported to improve computational reproducibility.

## 3. Results

### 3.1. Cohort characteristics

The MIMIC-IV cohort included 3,729 eligible intensive care stays, of which 1,027 (27.5%) resulted in in-hospital death. The cohort was divided into a training set of 2,983 stays, including 822 deaths, and an internal validation set of 746 stays, including 205 deaths. The independently constructed eICU-CRD external validation cohort included 611 stays, of which 188 (30.8%) resulted in in-hospital death.

Baseline characteristics are summarized in Table 1. Median age was 67 years (interquartile range, 59–76) in MIMIC-IV and 66 years (57–75) in eICU-CRD. The eICU-CRD cohort included a higher proportion of female patients and emergency admissions. Chronic obstructive pulmonary disease, diabetes, and metastatic cancer were recorded less frequently in eICU-CRD. The particularly large differences in recorded diabetes and metastatic cancer likely reflect cross-database variation in diagnosis coding and documentation, in addition to true case-mix differences. Hemoglobin was lower externally, and platelet-count categories differed substantially between cohorts.

**Table 1:**
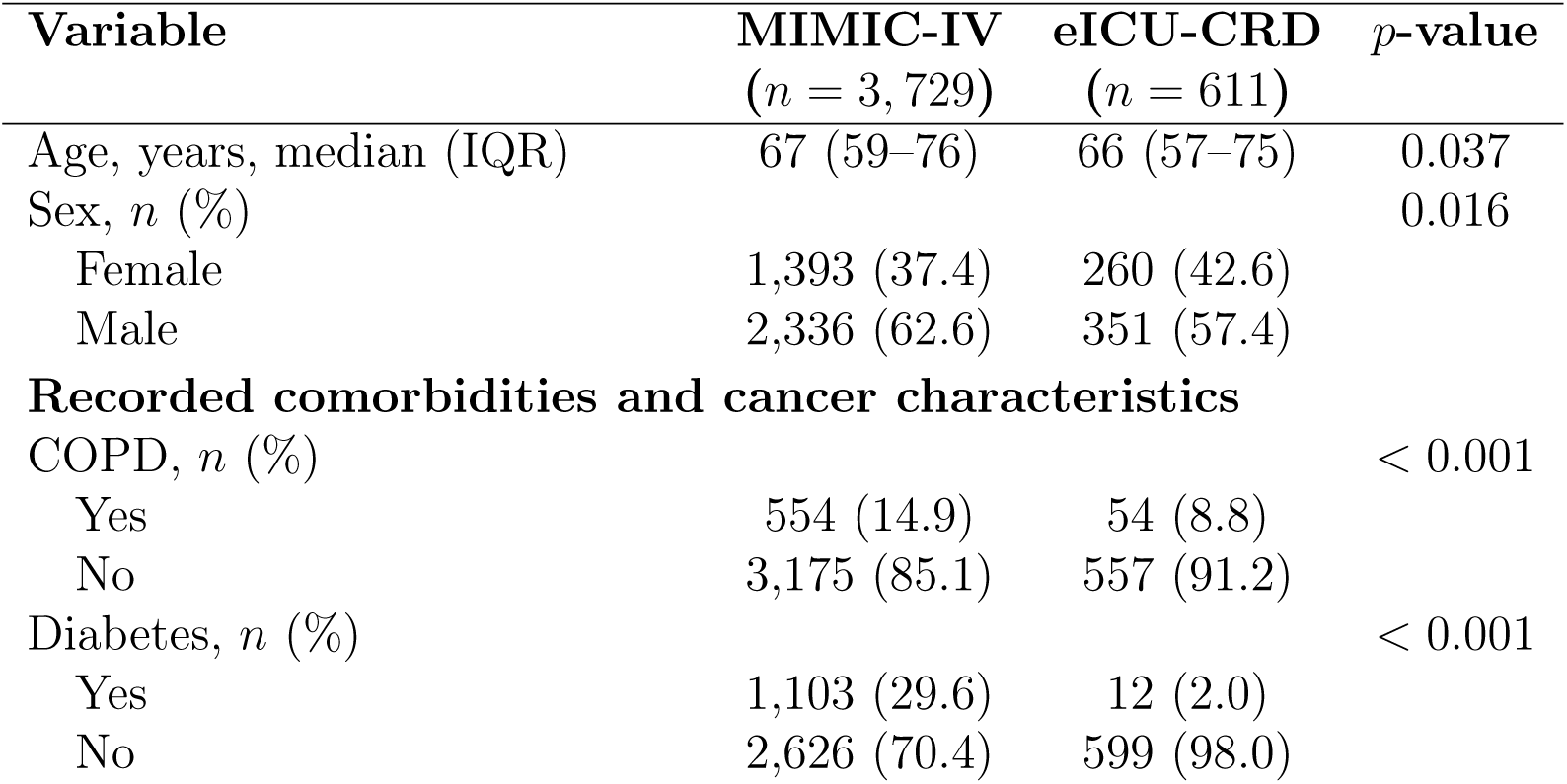
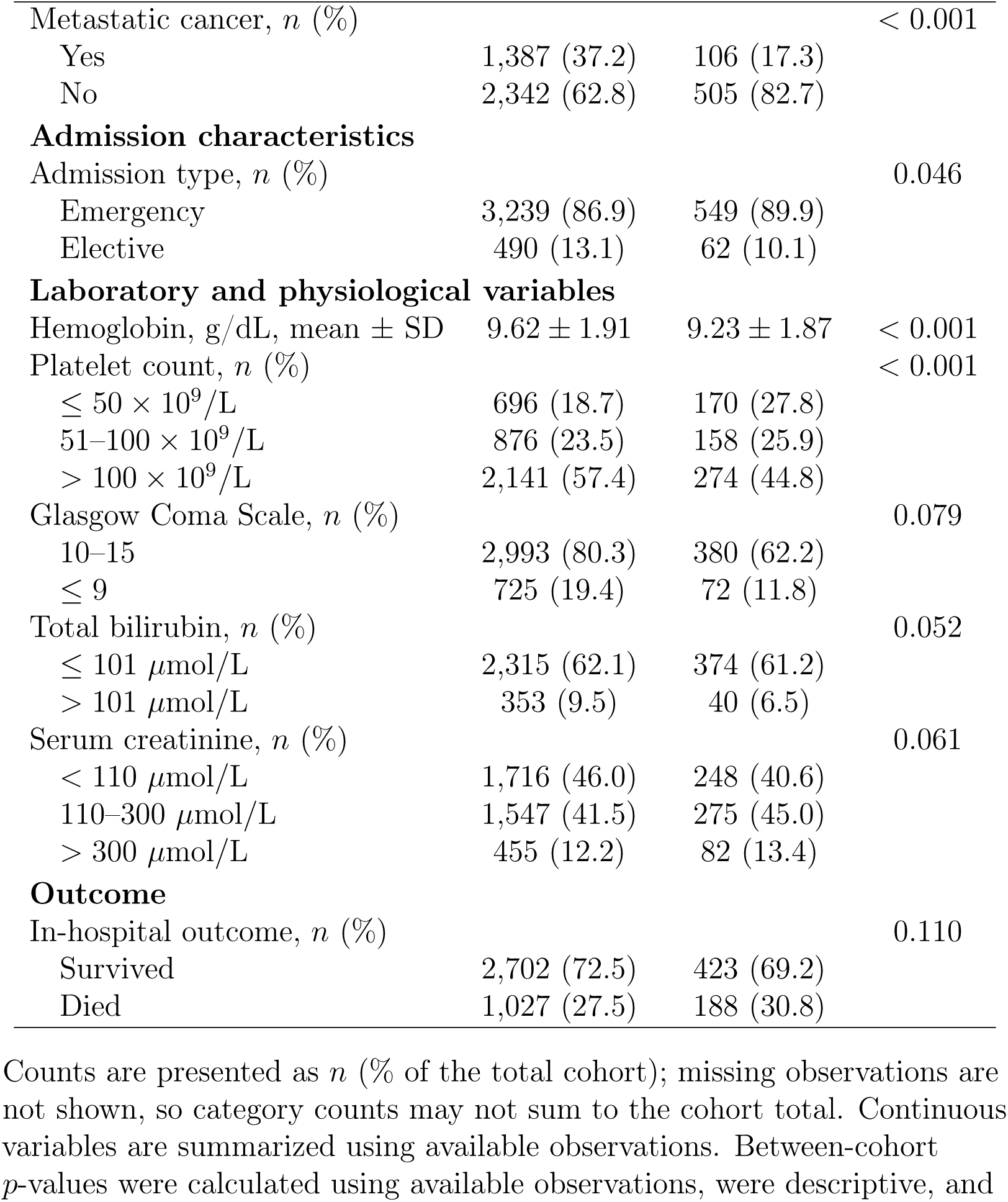
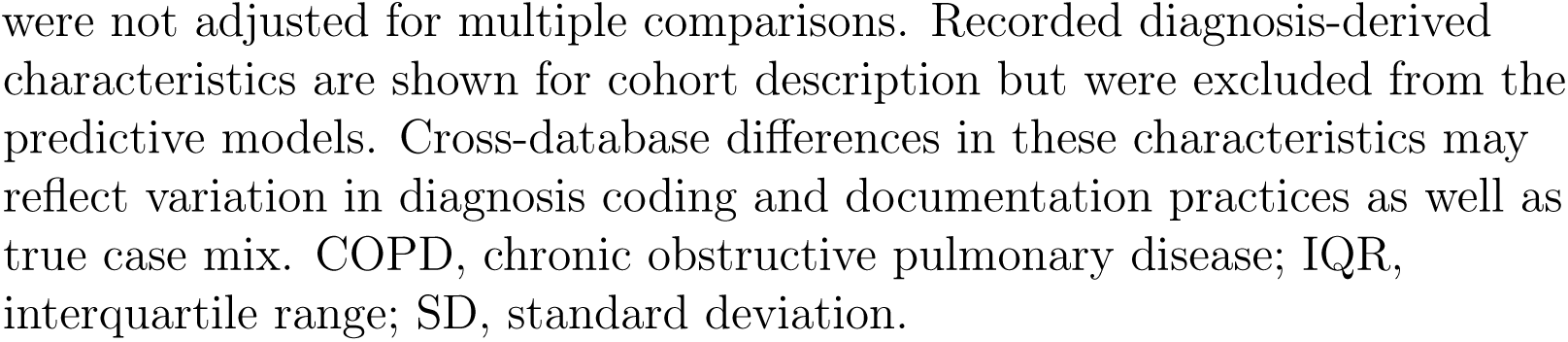
Baseline characteristics of the MIMIC-IV development cohort and the eICU-CRD external validation cohort.

Complete first-day Glasgow Coma Scale component data were available for 3,718 of 3,729 MIMIC-IV stays and 452 of 611 eICU-CRD stays. Among available observations, the distribution of the summarized Glasgow Coma Scale category did not differ substantially between cohorts. Hospital mortality was numerically higher in eICU-CRD than in MIMIC-IV (30.8% versus 27.5%), although the descriptive between-cohort comparison was not statistically significant (*p* = 0.110). Mean feature-level missingness was 36.6% in eICU-CRD compared with 20.2% in MIMIC-IV, and 39 of the 345 development predictors were entirely missing externally.

### 3.2. Final predictor set

After removal of identifiers, leakage-related variables, and same-admission diagnosis-derived variables, the initial modeling matrix contained 2,702 candidate predictors. Train-only missingness filtering excluded 2,358 predictors; no additional variables were removed by the all-missing or zero-variance filters. This left 344 retained predictors, and addition of the grouped Emergency-versus-Elective admission indicator produced a final modeling set of 345 predictors.

The final feature set comprised 10 demographic and admission variables, 39 first-24-hour vital-sign and Glasgow Coma Scale summaries, and 296 laboratory-related variables, including 288 item-based laboratory summaries and eight organ-dysfunction laboratory extrema or indicators (Table 2). The same fixed predictor set was used for hyperparameter tuning, final model fitting, held-out internal evaluation, SHAP analysis, calibration assessment, and locked external validation.

**Table 2:**
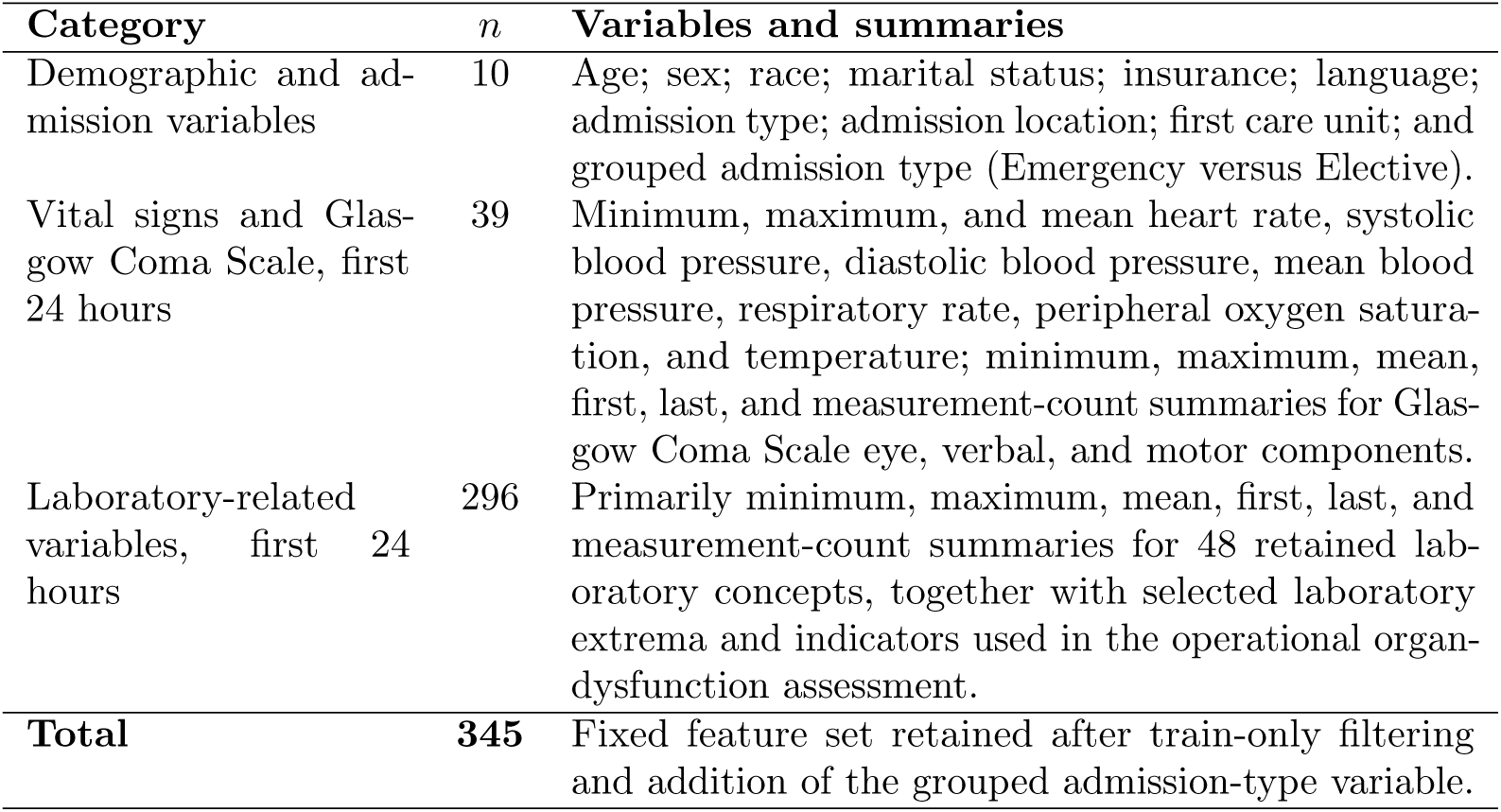
Predictor categories included in the final modeling set.

### 3.3. Internal validation performance

Fourteen predictive models were evaluated on the held-out internal validation cohort of 746 stays, including 205 in-hospital deaths. The most-frequent dummy classifier served as a non-informative reference and was not included in the comparative performance table.

GradientBoosting achieved the numerically highest internal validation AUROC of 0.8480 (95% confidence interval, 0.8173–0.8755), with an AUPRC of 0.6984 and a Brier score of 0.1346 (Table 3). HistGradientBoosting produced nearly identical discrimination, with an AUROC of 0.8477, an AUPRC of 0.6943, and a Brier score of 0.1350. Penalized logistic regression models achieved AUROC values of approximately 0.835–0.839. The confidence intervals of the leading models overlapped substantially, and the ranking should not be interpreted as evidence of definitive algorithmic superiority.

**Table 3:** Discrimination and probabilistic performance of machine learning models in the MIMIC-IV internal validation cohort.

| Model | AUROC (95% CI) | AUPRC | Brier score |
| --- | --- | --- | --- |
| **GradientBoosting** | **0.8480 (0.8173–0.8755)** | **0.6984** | **0.1346** |
| HistGradientBoosting | 0.8477 (0.8181–0.8759) | 0.6943 | 0.1350 |
| Elastic-net logistic regression | 0.8389 (0.8070–0.8696) | 0.6795 | 0.1386 |
| LASSO logistic regression | 0.8389 (0.8074–0.8684) | 0.6793 | 0.1386 |
| Ridge logistic regression | 0.8361 (0.8023–0.8666) | 0.6750 | 0.1400 |
| Balanced ridge logistic regression | 0.8348 (0.8009–0.8663) | 0.6725 | 0.1664 |
| Balanced LASSO logistic regression | 0.8345 (0.8026–0.8647) | 0.6685 | 0.1680 |
| AdaBoost | 0.8310 (0.7996–0.8596) | 0.6689 | 0.1933 |
| Random forest | 0.8256 (0.7914–0.8559) | 0.6544 | 0.1515 |
| Multilayer perceptron | 0.8215 (0.7862–0.8543) | 0.6526 | 0.1753 |
| ExtraTrees | 0.8214 (0.7890–0.8527) | 0.6493 | 0.1604 |
| $k$-nearest neighbors | 0.7702 (0.7294–0.8074) | 0.5760 | 0.1721 |
| Decision tree | 0.7646 (0.7284–0.8021) | 0.5387 | 0.2024 |
| Gaussian naive Bayes | 0.7341 (0.6945–0.7686) | 0.4595 | 0.3525 |

GradientBoosting was designated as the primary model because it had the highest prespecified internal validation AUROC. Its cross-validated training AUROC was 0.8355. At the operating threshold of 0.2706 selected from training-set out-of-fold predictions, GradientBoosting achieved a sensitivity of 0.7659, specificity of 0.7689, precision of 0.5567, F1 score of 0.6448, balanced accuracy of 0.7674, and accuracy of 0.7681 (Supplementary Table S1).

Receiver operating characteristic and precision–recall curves are shown in Figures 2 and 3. Confusion matrices calculated using training-derived operating thresholds are shown in Figure 4.

**Figure 2:**
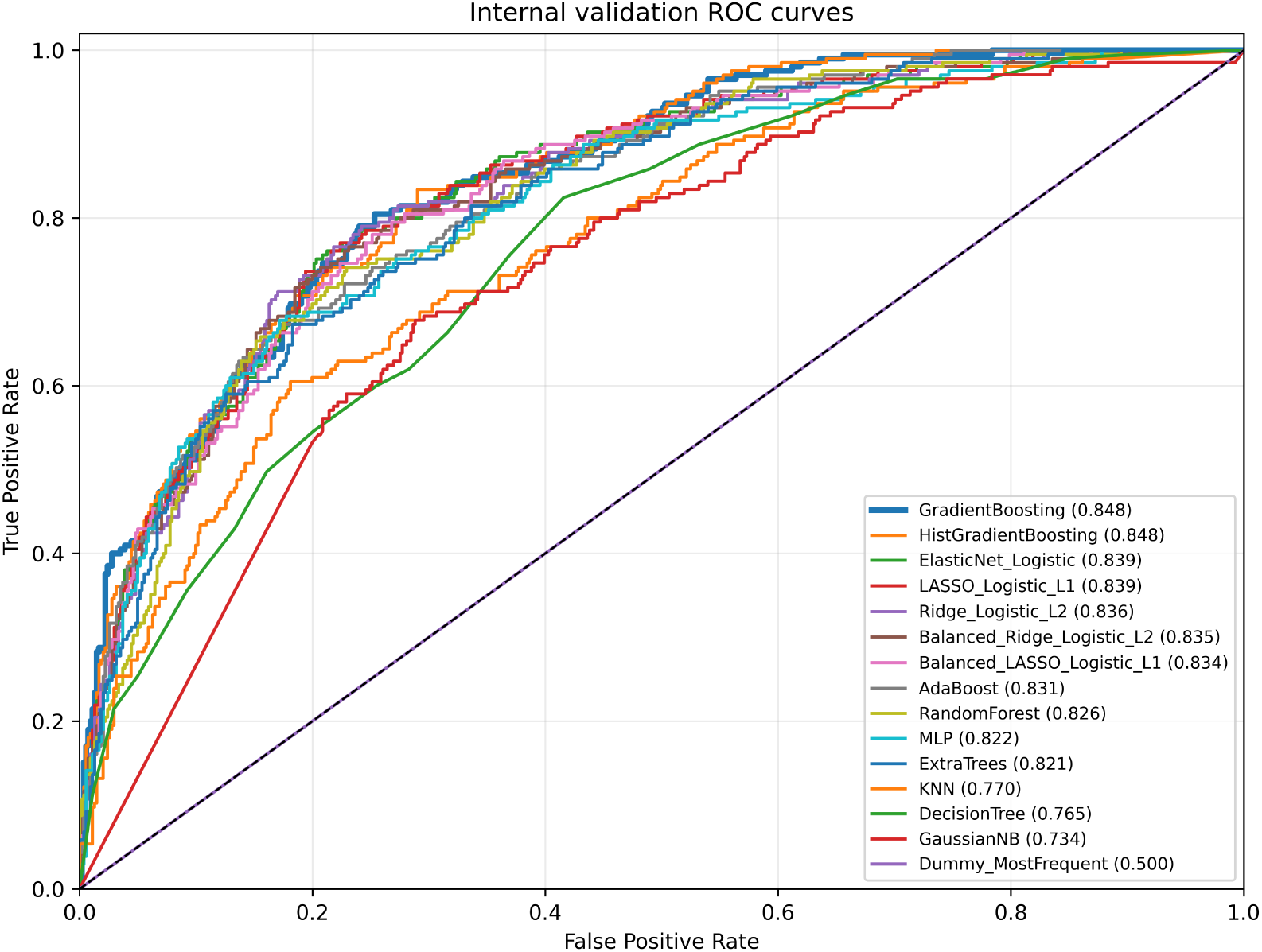
Receiver operating characteristic curves for evaluated models and the dummy baseline in the MIMIC-IV internal validation cohort. Curves show discrimination for the evaluated predictive models using the fixed 345-predictor feature set. GradientBoosting achieved the numerically highest internal validation AUROC.

**Figure 3:**
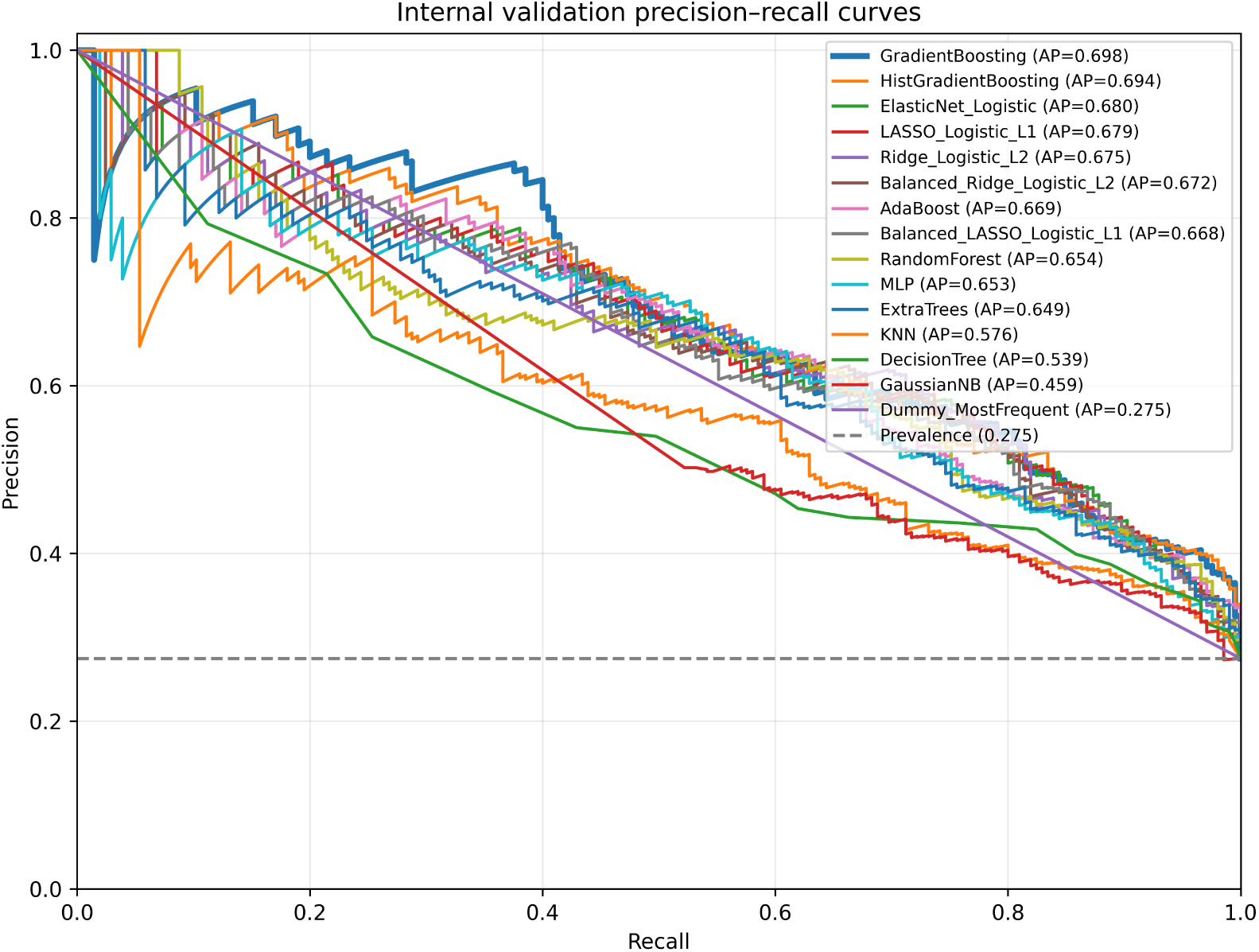
Precision–recall curves for evaluated models and the dummy baseline in the MIMIC-IV internal validation cohort. The internal validation mortality prevalence was 27.5%. GradientBoosting and HistGradientBoosting achieved the highest areas under the precision–recall curve.

**Figure 4:**
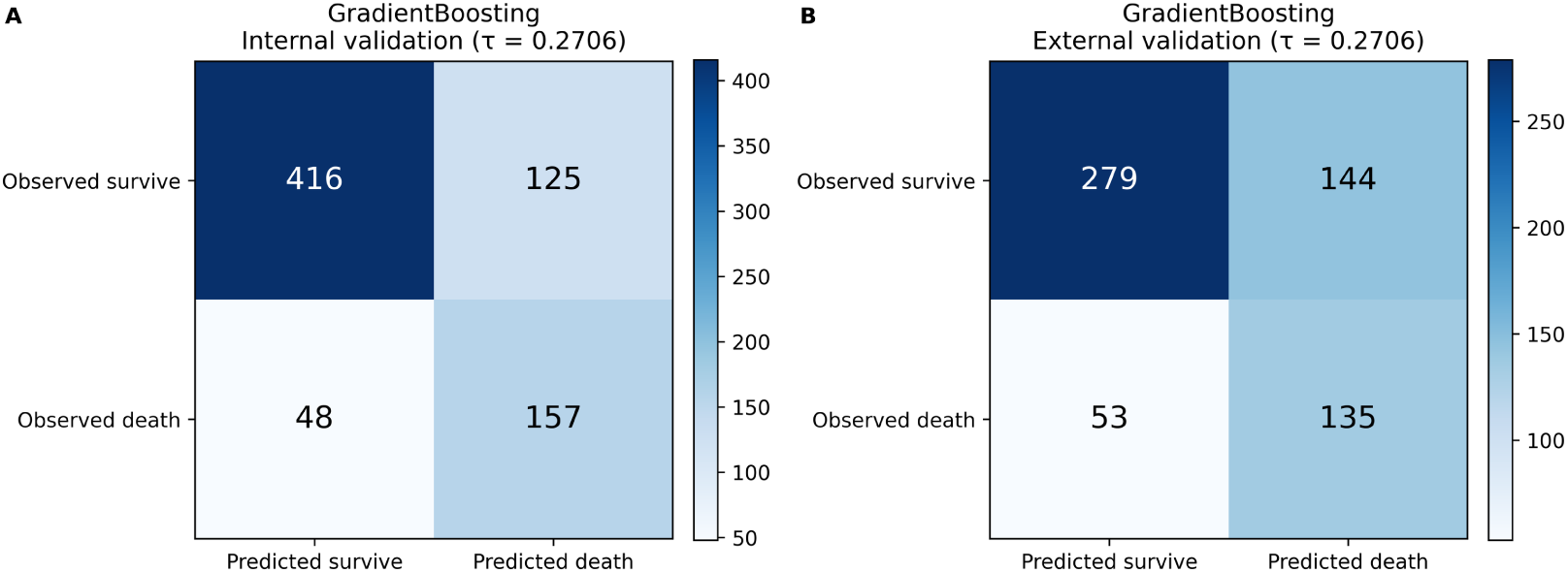
Confusion matrices for the primary GradientBoosting model in internal and external validation. Predicted classes were determined using the operating threshold of 0.2706 selected by maximizing Youden’s index using MIMIC-IV training-set out-of-fold predicted probabilities. The same locked threshold was used in eICU-CRD.

### 3.4. Model interpretation

SHAP analysis was performed for GradientBoosting, the prespecified primary model selected according to internal validation AUROC. Predictors were ranked according to their mean absolute SHAP values in the stratified internal validation subsample (Figure 5).

**Figure 5:**
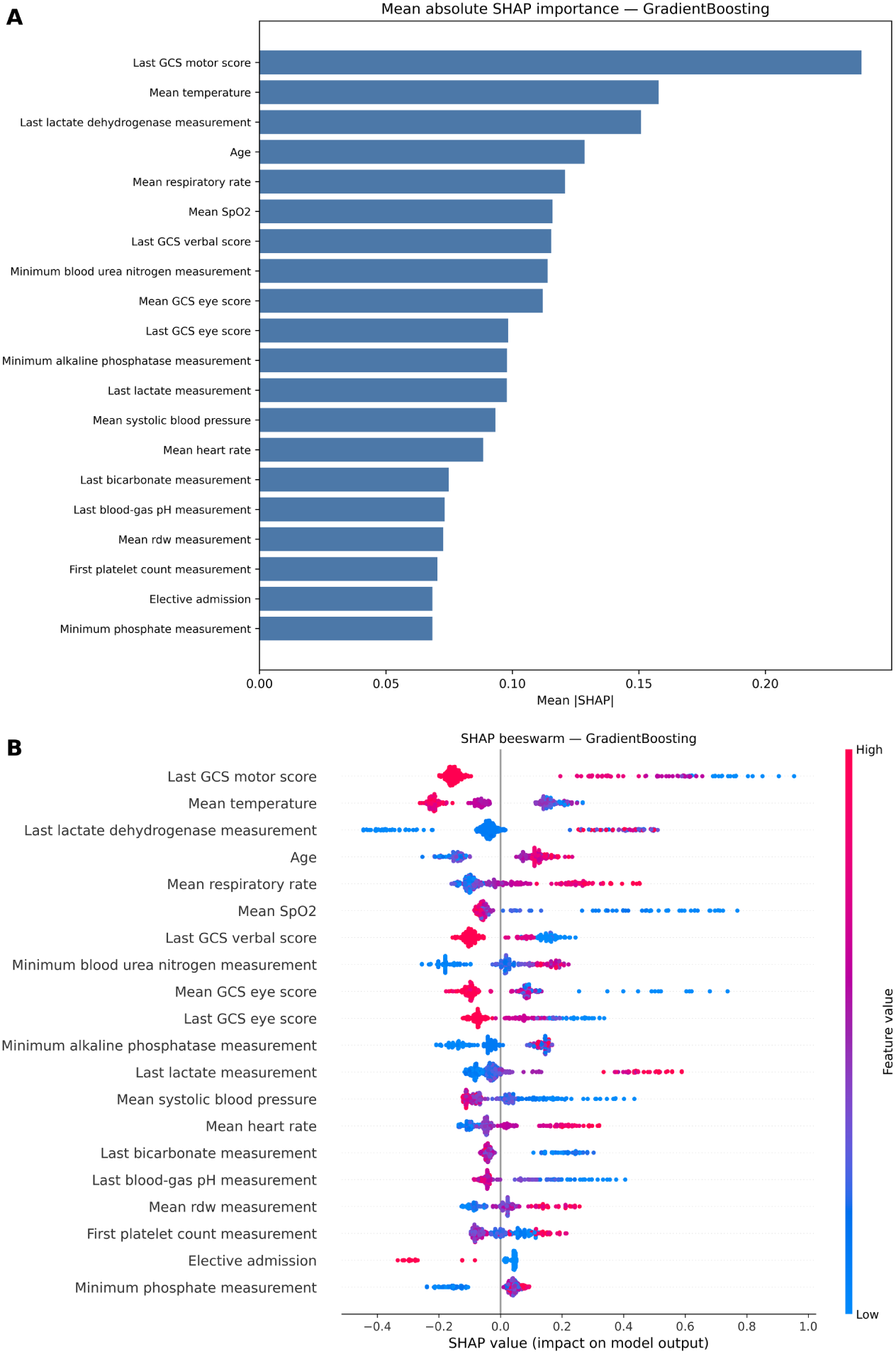
SHAP-based interpretation of the GradientBoosting model in the MIMIC-IV internal validation cohort. The figure presents the 20 predictors with the highest mean absolute SHAP values and the distribution and direction of their contributions. The analysis used a stratified background sample of 100 training observations and a stratified explanation sample of 300 internal validation observations. SHAP, SHapley Additive exPlanations.

The feature with the highest global SHAP importance was the last recorded Glasgow Coma Scale motor score during the first 24 hours (mean absolute SHAP value, 0.238). This was followed by mean temperature (0.158), the last lactate dehydrogenase measurement (0.151), age (0.129), mean respiratory rate (0.121), mean peripheral oxygen saturation (0.116), the last Glasgow Coma Scale verbal score (0.115), minimum blood urea nitrogen (0.114), mean Glasgow Coma Scale eye score (0.112), and the last Glasgow Coma Scale eye score (0.098).

Other predictors among the 20 most influential features included minimum alkaline phosphatase, last serum lactate, mean systolic blood pressure, mean heart rate, last bicarbonate, last blood-gas pH, mean red-cell distribution width, first platelet count, elective admission status, and minimum phosphate. The ranking indicated that the model relied primarily on neurological status, temperature, respiratory physiology, oxygenation, laboratory markers of organ dysfunction, age, and admission context. SHAP values quantify contributions to fitted predictions and should not be interpreted as causal effects or independent clinical risk factors.

### 3.5. Probabilistic performance and calibration

Reliability curves with 10 quantile-based bins were used to examine agreement between predicted and observed mortality probabilities (Figure 6). Overall probabilistic prediction error was additionally summarized using the Brier score.

**Figure 6:**
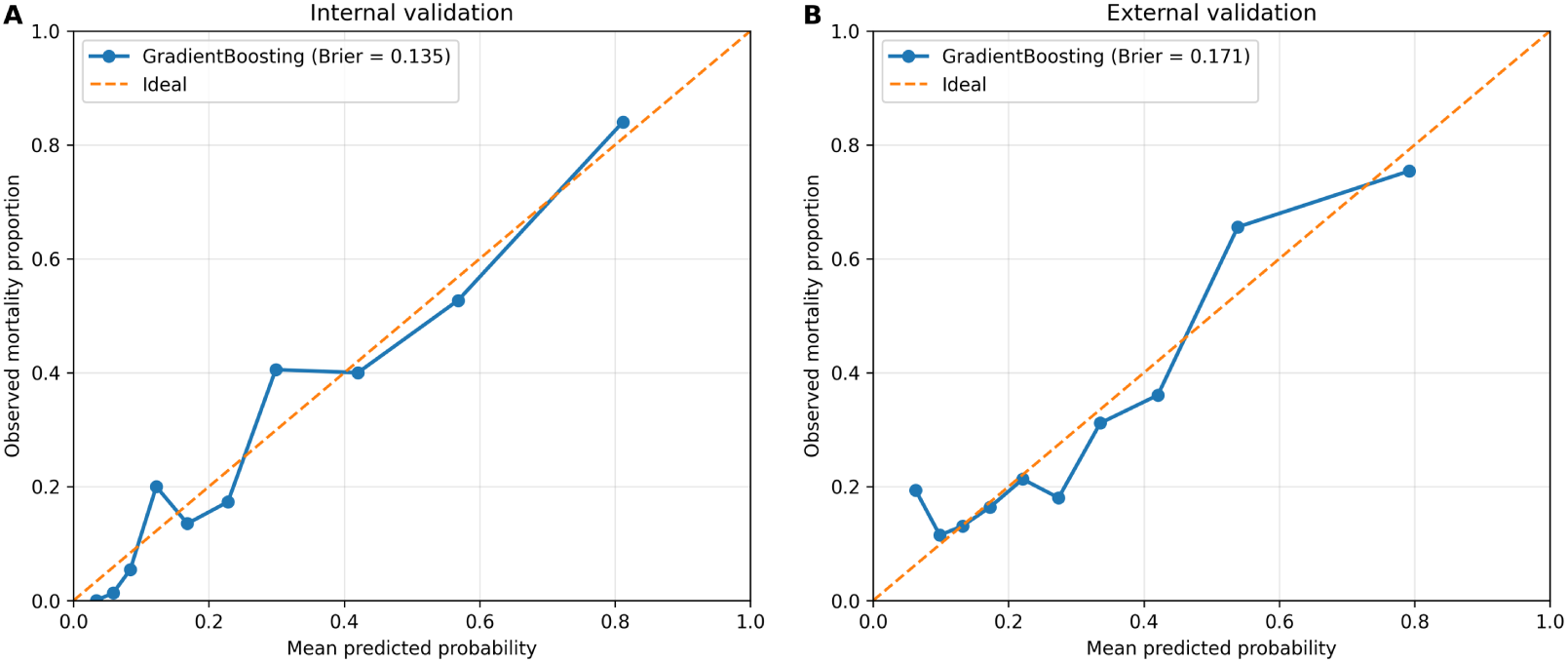
Calibration of the primary GradientBoosting model in internal and external validation. Predicted probabilities were divided into 10 quantile-based bins. The diagonal line represents perfect agreement between predicted and observed mortality probabilities. The model was applied to eICU-CRD without recalibration.

GradientBoosting had the lowest internal validation Brier score among the evaluated models (0.1346), closely followed by HistGradientBoosting (0.1350), elastic-net and LASSO logistic regression (both 0.1386), and ridge logistic regression (0.1400). The GradientBoosting Brier score increased to 0.1709 under external validation, indicating deterioration in overall probabilistic performance after transport to eICU-CRD. Reliability curves and Brier scores were interpreted jointly with discrimination and were not treated as evidence of complete calibration.

### 3.6. External validation

The locked MIMIC-IV pipelines were applied to the eICU-CRD external validation cohort of 611 stays, including 188 in-hospital deaths. The fixed 345-predictor feature set, training-fitted preprocessing parameters, selected hyperparameters, and model-specific operating thresholds derived from MIMIC-IV training-set out-of-fold predictions were used without modification. After final concept and unit harmonization, 39 of the 345 development predictors were entirely missing in eICU-CRD. Mean feature-level missingness was 36.6% externally and 20.2% in MIMIC-IV. Complete first-day Glasgow Coma Scale eye, verbal, and motor component data were available for 452 external stays. External performance is summarized in Table 4.

**Table 4:**
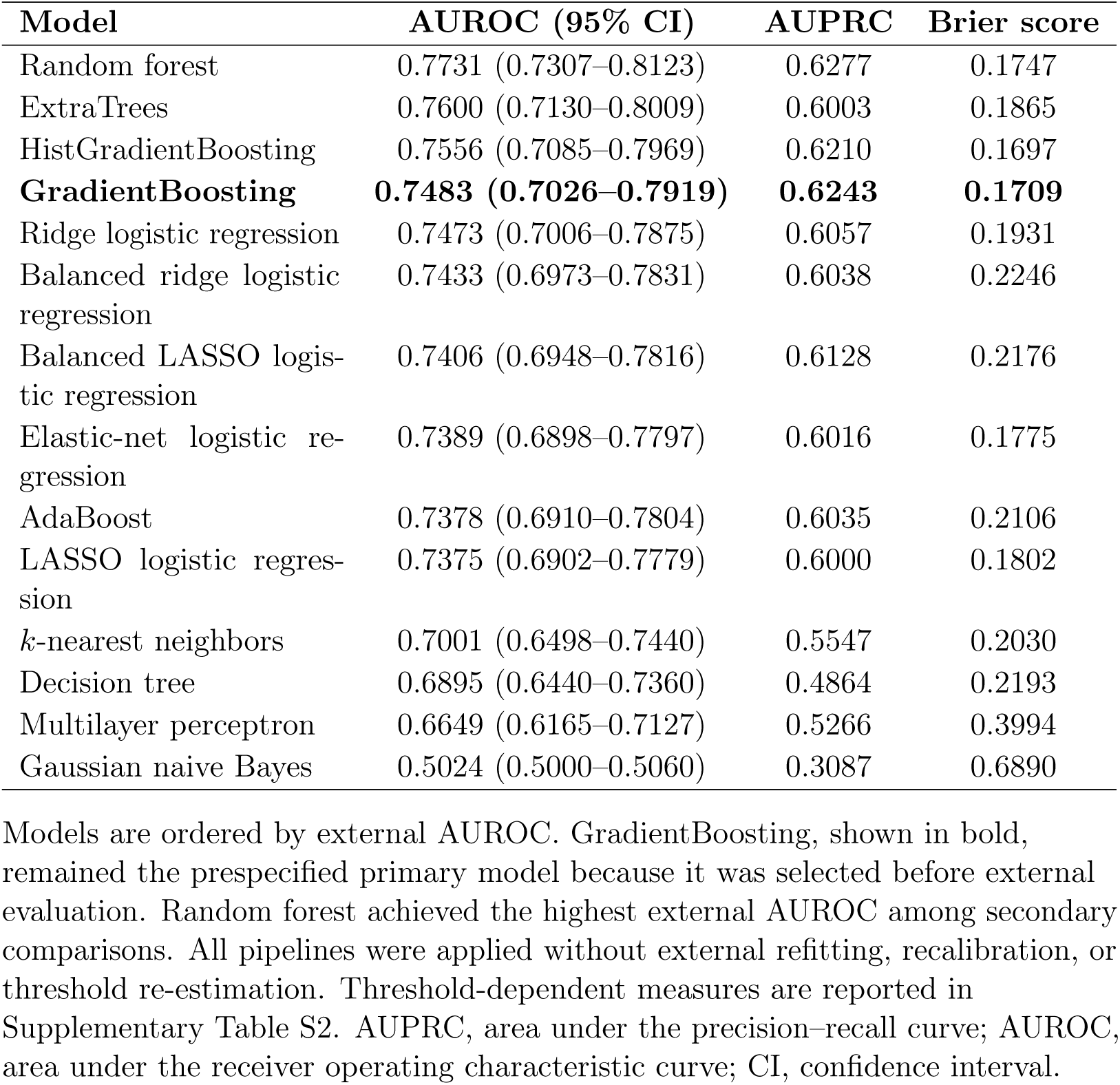
Discrimination and probabilistic performance under locked external validation in eICU-CRD.

| Model | AUROC (95% CI) | AUPRC | Brier score |
| --- | --- | --- | --- |
| Random forest | 0.7731 (0.7307–0.8123) | 0.6277 | 0.1747 |
| ExtraTrees | 0.7600 (0.7130–0.8009) | 0.6003 | 0.1865 |
| HistGradientBoosting | 0.7556 (0.7085–0.7969) | 0.6210 | 0.1697 |
| **GradientBoosting** | **0.7483 (0.7026–0.7919)** | **0.6243** | **0.1709** |
| Ridge logistic regression | 0.7473 (0.7006–0.7875) | 0.6057 | 0.1931 |
| Balanced ridge logistic regression | 0.7433 (0.6973–0.7831) | 0.6038 | 0.2246 |
| Balanced LASSO logistic regression | 0.7406 (0.6948–0.7816) | 0.6128 | 0.2176 |
| Elastic-net logistic regression | 0.7389 (0.6898–0.7797) | 0.6016 | 0.1775 |
| AdaBoost | 0.7378 (0.6910–0.7804) | 0.6035 | 0.2106 |
| LASSO logistic regression | 0.7375 (0.6902–0.7779) | 0.6000 | 0.1802 |
| $k$-nearest neighbors | 0.7001 (0.6498–0.7440) | 0.5547 | 0.2030 |
| Decision tree | 0.6895 (0.6440–0.7360) | 0.4864 | 0.2193 |
| Multilayer perceptron | 0.6649 (0.6165–0.7127) | 0.5266 | 0.3994 |
| Gaussian naive Bayes | 0.5024 (0.5000–0.5060) | 0.3087 | 0.6890 |

The prespecified primary GradientBoosting model achieved an external AUROC of 0.7483 (95% confidence interval, 0.7026–0.7919), compared with an internal validation AUROC of 0.8480. Its external AUPRC was 0.6243, and its Brier score increased from 0.1346 internally to 0.1709 externally. At the locked MIMIC-IV operating threshold of 0.2706, GradientBoosting achieved a sensitivity of 0.7181, specificity of 0.6596, precision of 0.4839, F1 score of 0.5782, balanced accuracy of 0.6888, and accuracy of 0.6776 (Supplementary Table S2).

Random forest showed the numerically highest external discrimination, with an AUROC of 0.7731 (95% confidence interval, 0.7307–0.8123), AUPRC of 0.6277, and Brier score of 0.1747. ExtraTrees and HistGradientBoosting achieved external AUROCs of 0.7600 and 0.7556, respectively. The confidence intervals of the leading tree-ensemble models overlapped.

The higher external AUROC of random forest was treated as a secondary comparative finding. GradientBoosting remained the primary model because it had been selected before external evaluation according to the prespecified internal validation criterion. Retrospectively replacing it with the best-performing external model would use eICU-CRD for model selection and would no longer constitute independent validation.

The external performance decline occurred despite harmonization of first-day Glasgow Coma Scale and laboratory variables. Residual differences in case mix, documentation, measurement frequency, predictor availability, and care processes remained substantial. The findings indicate that the MIMIC-IV models retained moderate discriminative information under locked external validation but did not demonstrate fully stable transportability across databases. Receiver operating characteristic and precision–recall curves for all evaluated models under locked external validation are provided in Supplementary Figure S1 and Supplementary Figure S2, respectively.

## 4. Discussion

### 4.1. Principal findings

In this study, we developed and externally evaluated machine learning models for predicting subsequent in-hospital mortality among intensive care unit patients with cancer and sepsis who remained in intensive care through a 24-hour prediction landmark. The development cohort included 3,729 MIMIC-IV stays, and locked external validation was performed in 611 eICU-CRD stays. Fourteen predictive algorithms were compared under a shared preprocessing and tuning framework after excluding same-admission diagnosis-derived variables whose availability by the prediction landmark could not be established. GradientBoosting was selected as the primary model according to the prespecified internal validation AUROC criterion.

GradientBoosting achieved an internal validation AUROC of 0.8480, an AUPRC of 0.6984, and a Brier score of 0.1346. HistGradientBoosting produced nearly identical internal discrimination and probabilistic performance, and the confidence intervals of the leading models overlapped substantially. These findings support the ability of both boosting methods and regularized linear models to extract prognostic information from routinely collected first-day intensive care data, but they do not demonstrate clear superiority of one algorithm over all alternatives. The relatively small performance differences among the leading models are also consistent with evidence that more complex machine learning methods do not invariably outperform well-specified regression models in clinical prediction tasks [16].

SHAP analysis indicated that the primary model relied on a clinically coherent combination of neurological status, temperature, respiratory physiology, oxygenation, age, and laboratory measurements. The last Glasgow Coma Scale motor score was the most influential feature, followed by mean temperature, the last lactate dehydrogenase measurement, age, mean respiratory rate, mean peripheral oxygen saturation, additional Glasgow Coma Scale components, blood urea nitrogen, and serum lactate. These variables plausibly reflect acute neurological and physiological derangement among critically ill patients. Nevertheless, SHAP values describe contributions to fitted predictions rather than causal effects, independent risk factors, or treatment targets, and they should not be interpreted as establishing biological mechanisms or clinical actionability [28].

The principal GradientBoosting model retained moderate discrimination under locked external validation, with an AUROC of 0.7483 and an AUPRC of 0.6243 in eICU-CRD. However, its AUROC declined from 0.8480 internally, and its Brier score increased from 0.1346 to 0.1709. At the training-derived operating threshold, sensitivity was 0.7181, specificity was 0.6596, and precision was 0.4839. These changes indicate that both probability estimates and threshold-dependent classification performance were affected by transport to a different database. Such deterioration is consistent with prediction-model methodology showing that discrimination, calibration, and operating characteristics may change across populations with different case mix, measurement practices, documentation systems, and care processes [19, 20, 21].

Model ranking also changed externally. Random forest achieved the highest external AUROC of 0.7731, whereas its internal validation AUROC was lower than that of the two boosting models and several penalized logistic regression models. Because random forest had not been selected as the primary model before external evaluation, this result was treated as a secondary comparison rather than a basis for replacing GradientBoosting. The change in ranking illustrates that the model with the highest discrimination in a development environment may not be the most robust under dataset shift. It also reinforces the importance of retaining external data for evaluation rather than using them for post hoc model selection.

Differences between the development and external cohorts provide plausible explanations for the performance decline. The eICU-CRD cohort had a somewhat higher mortality rate, different admission and recorded comorbidity distributions, and substantially greater feature missingness. Thirty-nine of the 345 predictors were entirely missing externally, and mean feature-level missingness was 36.6% in eICU-CRD compared with 20.2% in MIMIC-IV. Complete first-day Glasgow Coma Scale component data were available for approximately 74% of external stays. Although major laboratory concepts and units were harmonized before locked scoring, residual differences in measurement frequency, clinical documentation, institutional workflows, and the meaning of unavailable variables likely remained. The external results should therefore be interpreted as evidence of incomplete transportability rather than failure of a single algorithm.

From a clinical perspective, the models may support identification of patients who remain at elevated risk after the first 24 hours of intensive care. Potential applications could include prioritization of reassessment, monitoring, or multidisciplinary discussion. However, the models are not admission-time tools and should not be used as standalone systems for treatment limitation, resource allocation, or other high-stakes decisions. Clinical use would require prospective validation, local assessment of calibration and operating thresholds, evaluation of net benefit, and integration with clinician judgment.

### 4.2. Comparison with previous studies

Published prediction studies focused specifically on critically ill patients with both cancer and sepsis remain limited. Yuan et al. developed a LASSO-selected logistic regression nomogram for hospital mortality among patients with sepsis and solid cancer using MIMIC-IV and externally validated the model in eICU-CRD [9]. Their reported AUROC was 0.726 in internal validation and 0.756 in external validation. Tang et al. subsequently evaluated interpretable machine learning models in patients with lung cancer and sepsis and also included external validation [10]. These studies demonstrate increasing interest in malignancy-specific sepsis prediction but were restricted to solid cancer or a single malignancy type.

The present study extends this literature in several ways. It included both solid and hematologic malignancies, compared a broader range of linear, tree-based, boosting, instance-based, probabilistic, and neural-network models, and evaluated discrimination, precision–recall performance, probabilistic prediction error, threshold-dependent classification, SHAP-based interpretation, and locked external performance. Same-admission diagnosis-derived predictors were excluded from modeling because their availability at the 24-hour landmark could not be established consistently, reducing the risk that discharge-level coding information would inflate model performance.

The primary GradientBoosting model achieved a higher internal AUROC than the nomogram reported by Yuan et al. (0.8480 versus 0.726), but its external AUROC of 0.7483 was similar to the previously reported value of 0.756 [9]. Random forest achieved an external AUROC of 0.7731 in the present analysis, although this was a secondary model comparison. These values should not be interpreted as a direct head-to-head comparison because the studies differed in cohort definitions, cancer populations, predictor availability, preprocessing, and outcome ascertainment. More importantly, the smaller difference in external than internal performance suggests that improvements observed within one development database may not fully persist after transport.

Studies in broader sepsis populations have also reported favorable performance for gradient-boosted trees and other machine learning approaches. Hou et al. reported that XGBoost outperformed logistic regression and SAPS II for 30-day mortality prediction in MIMIC-III [11]. Li et al. and Wang et al. developed interpretable machine learning models for sepsis mortality and identified neurological, physiological, and laboratory variables among important predictors [12, 13]. Bao et al. evaluated sepsis mortality models across MIMIC-IV and eICU-CRD, and Zhang et al. emphasized multicenter validation across heterogeneous clinical settings [14, 15]. The importance of neurological status, respiratory physiology, temperature, oxygenation, renal markers, and lactate-related measurements in the present study is broadly consistent with this literature.

A central contribution of the current analysis is the use of a locked external validation design. The fixed feature set, preprocessing parameters, fitted models, and operating thresholds were transferred to eICU-CRD without refitting, recalibration, feature reselection, or threshold optimization. This approach produced less favorable results than might be obtained after local model updating, but it provides a more informative estimate of transportability. External validation guidance similarly emphasizes evaluation in populations that differ meaningfully from the development data and separation of evaluation from subsequent model optimization [19, 20, 21].

### 4.3. Strengths and limitations

This study had several strengths. It used an updated version of MIMIC-IV for development and an independent multicenter database for external validation. Both solid and hematologic malignancies were represented. Fourteen predictive algorithms were evaluated under a shared feature set, preprocessing framework, and tuning procedure, reducing inconsistency across model comparisons. Feature filtering, preprocessing estimation, hyperparameter tuning, and operating-threshold selection were restricted to the development data. The external cohort was scored using frozen pipelines, and performance was evaluated using discrimination, precision–recall performance, threshold-dependent metrics, Brier scores, reliability curves, and model interpretation rather than AUROC alone. These choices are consistent with contemporary recommendations for clinical prediction-model evaluation and reporting [17, 25].

Several limitations should also be considered. First, the study was retrospective and relied on routinely collected electronic health record data. Missing measurements, recording errors, differences in sampling frequency, and changes in clinical practice may have affected both cohort construction and predictor quality. The observed associations may also reflect local documentation and testing practices rather than only patient physiology.

Second, the prediction landmark was 24 hours after intensive care unit admission. Patients who died, were discharged, or left intensive care before the landmark were not represented. The findings therefore apply to patients who remained in intensive care through the first 24 hours and should not be generalized to admission-time prediction or very early deterioration. The landmark design may also yield a clinically different population from studies that make predictions at or before intensive care admission.

Third, sepsis was defined using a database-compatible operational definition based on suspected infection and selected laboratory evidence of organ dysfunction rather than a complete Sequential Organ Failure Assessment-based implementation of Sepsis-3. Misclassification is therefore possible, and comparisons with studies using different sepsis definitions should be interpreted cautiously. Cancer status was also determined retrospectively using hospitalization-level diagnosis codes, although these codes were used only for cohort eligibility and not as model predictors.

Fourth, cross-database feature harmonization remained incomplete. Thirty-nine predictors were entirely missing in eICU-CRD, external feature-level missingness was substantially higher, and complete Glasgow Coma Scale components were available for only approximately three quarters of external stays. Although pH, red-cell distribution width, ionized calcium, and other major laboratory concepts were aligned by clinical meaning and unit, residual differences in item definitions, measurement frequency, assay methods, and institutional workflows may have influenced predictions. Such differences are recognized sources of reduced transportability across electronic health record systems [19, 20].

Fifth, same-admission diagnosis-derived variables were intentionally excluded because their availability by the 24-hour landmark could not be established. This decision reduced the risk of temporal leakage but also limited representation of chronic comorbidity, cancer subtype, metastatic burden, and other baseline disease characteristics. More reliable time-stamped pre-admission or early-admission data could improve case-mix adjustment in future studies. Cancer is also highly heterogeneous, and structured information on cancer stage, treatment status, neutropenia, recent chemotherapy, stem-cell transplantation, functional status, and goals of care was not comprehensively available. Outcomes among critically ill oncology patients are known to vary according to malignancy status, functional condition, and acute organ dysfunction [6, 8].

Sixth, a single held-out internal validation split was used after cross-validated hyperparameter tuning. The same internal validation cohort was also used to select the primary model according to AUROC and to report its internal performance. Although no model parameters were fitted on this cohort, selection and evaluation on the same split may introduce modest optimism into the primary internal estimate. Repeated resampling, nested validation, temporal validation, or evaluation in additional independent cohorts would provide more stable estimates of model uncertainty [21, 26].

Seventh, calibration was assessed using Brier scores and reliability curves, but formal calibration intercepts and slopes were not reported, and no external recalibration was performed. Decision-curve analysis was also not conducted. The locked thresholds were intentionally preserved to evaluate transportability, but they may not reflect appropriate clinical trade-offs in another institution. Future work should evaluate calibration-in-the-large, calibration slopes, local model updating, clinically selected thresholds, and net benefit across relevant risk ranges [18, 19].

Finally, predictive performance and SHAP-based interpretability do not establish clinical utility. A model with acceptable discrimination may still have inadequate calibration, unsuitable decision thresholds, or limited net benefit [17, 18]. Prospective multicenter studies are needed to determine whether predictions can be integrated into workflow, whether clinicians interpret them appropriately, and whether model-supported decisions improve monitoring, treatment, communication, or patient outcomes. Continuous monitoring would also be required to detect performance drift after implementation.

## 5. Conclusions

This study developed and externally evaluated machine learning models for predicting subsequent in-hospital mortality among intensive care unit patients with cancer and sepsis using demographic, admission, physiological, neurological, and laboratory information from the first 24 hours of intensive care. GradientBoosting achieved strong internal discrimination and retained moderate discrimination under locked external validation, although performance and probabilistic accuracy declined in eICU-CRD. Random forest achieved the highest external AUROC as a secondary comparison, illustrating that internal model ranking did not remain stable across databases.

The findings support the potential value of routinely collected first-day data for risk stratification among patients who remain in intensive care through the 24-hour landmark, while also demonstrating that favorable internal performance does not ensure stable transportability. The models should not be interpreted as admission-time predictors or standalone clinical decision systems. Prospective multicenter validation, local calibration and threshold assessment, and evaluation of clinical utility are required before implementation.

## CRediT authorship contribution statement

JS, SP, YS, MH, and NK contributed equally to this work. JS performed data curation, feature engineering, formal analysis, model development, validation, visualization, and preparation of the original manuscript draft. YS contributed to methodological review, validation of the analytical and reporting framework, interpretation of the results, journal-oriented restructuring, visualization, and substantial revision of the manuscript. SP, MH, and NK contributed to study development, methodological review, interpretation of the findings, and critical revision of the manuscript. GP and KA contributed to clinical interpretation of the findings and critical revision of the manuscript. MP conceived and supervised the study, coordinated the project, contributed to interpretation of the results, and critically revised the manuscript. All authors reviewed and approved the final manuscript.

## Funding

This research did not receive any specific grant from funding agencies in the public, commercial, or not-for-profit sectors.

## Declaration of competing interest

The authors declare that they have no known competing financial interests or personal relationships that could have appeared to influence the work reported in this paper.

## Ethics approval and consent to participate

This study used de-identified data from MIMIC-IV version 3.1 and the eICU Collaborative Research Database version 2.0. Access was obtained through PhysioNet after completion of the required credentialing, human-subjects research training, and applicable data-use agreements. Because the study involved secondary analysis of de-identified records and no direct interaction with participants, no additional institutional review board approval or individual informed consent was required.

## Data availability

The MIMIC-IV version 3.1 and eICU Collaborative Research Database version 2.0 data analyzed in this study are available to credentialed researchers through PhysioNet, subject to completion of the required research training and data-use agreements. Because these are third-party controlled-access datasets, patient-level records and derived patient-level extracts cannot be redistributed by the authors. No new patient data were collected.

## Code availability

The analysis code supporting the findings of this study may be obtained from the corresponding author upon reasonable request, subject to the applicable PhysioNet data-use requirements.

## Acknowledgments

The authors have no additional acknowledgments.

## Declaration of generative AI and AI-assisted technologies in the manuscript preparation process

During the preparation of this work, the authors used OpenAI ChatGPT for editorial restructuring, language refinement, and drafting assistance, and Anthropic Claude for language refinement. After using these tools, the authors reviewed and edited the content as needed, verified all numerical and methodological statements against the original analyses and source materials, and take full responsibility for the content of the publication. The tools were not used to generate or modify patient data, construct study cohorts, perform statistical analyses, train or evaluate the machine learning models, or generate the reported results.

## Notes

### Competing Interest Statement

The authors have declared no competing interest.

## References

[1] H. Sung, J. Ferlay, R. L. Siegel, M. Laversanne, I. Soerjomataram, A. Jemal, F. Bray, Global cancer statistics 2020: GLOBOCAN estimates of incidence and mortality worldwide for 36 cancers in 185 countries, CA: A Cancer Journal for Clinicians 71 (3) (2021) 209–249. doi:10.3322/caac.21660.

[2] M. Singer, C. S. Deutschman, C. W. Seymour, M. Shankar-Hari, D. Annane, M. Bauer, R. Bellomo, G. R. Bernard, J.-D. Chiche, C. M. Coopersmith, R. S. Hotchkiss, M. M. Levy, J. C. Marshall, G. S. Martin, S. M. Opal, G. D. Rubenfeld, T. van der Poll, J.-L. Vincent, D. C. Angus, The third international consensus definitions for sepsis and septic shock (Sepsis-3), JAMA 315 (8) (2016) 801–810. doi:10.1001/jama.2016.0287.

[3] M. D. Williams, L. A. Braun, L. M. Cooper, J. Johnston, R. V. Weiss, R. L. Qualy, W. Linde-Zwirble, Hospitalized cancer patients with severe sepsis: Analysis of incidence, mortality, and associated costs of care, Critical Care 8 (5) (2004) R291–R298. doi:10.1186/cc2893.

[4] L. Nazer, M. A. Lopez-Olivo, J. A. Cuenca, W. Awad, A. R. Brown, A. Abusara, M. Sirimaturos, R. S. Hicklen, J. L. Nates, All-cause mortality in cancer patients treated for sepsis in intensive care units: A systematic review and meta-analysis, Supportive Care in Cancer 30 (12) (2022) 10099–10109. doi:10.1007/s00520-022-07392-w.

[5] L. Edwards, E. Nelmes, M. Ardissino, H. L. J. Qi, S. Jhanji, D. B. Antcliffe, K. C. Tatham, The evolution of mortality from sepsis in patients with cancer: A systematic review and meta-analysis, Journal of the Intensive Care Society 27 (1) (2026) 55–65. doi:10.1177/17511437251363762.

[6] M. Soares, P. Caruso, E. Silva, J. M. M. Teles, S. M. A. Lobo, G. Friedman, F. Dal Pizzol, P. V. C. Mello, F. A. Bozza, U. V. A. Silva, et al., Characteristics and outcomes of patients with cancer requiring admission to intensive care units: A prospective multicenter study, Critical Care Medicine 38 (1) (2010) 9–15. doi:10.1097/CCM.0b013e3181c0349e.

[7] P. Schellongowski, M. Benesch, T. Lang, F. Traunmuller, C. Zauner, K. Laczika, G. J. Locker, M. Frass, T. Staudinger, Comparison of three severity scores for critically ill cancer patients, Intensive Care Medicine 30 (3) (2004) 430–436. doi:10.1007/s00134-003-2043-1.

[8] M. Soares, U. V. A. Silva, J. M. M. Teles, E. Silva, P. Caruso, S. M. A. Lobo, F. Dal Pizzol, L. P. Azevedo, F. B. de Carvalho, J. I. F. Salluh, Validation of four prognostic scores in patients with cancer admitted to brazilian intensive care units: Results from a prospective multicenter study, Intensive Care Medicine 36 (7) (2010) 1188–1195. doi:10.1007/s00134-010-1807-7.

[9] Z.-N. Yuan, Y.-J. Xue, H.-J. Wang, S.-N. Qu, C.-L. Huang, H. Wang, H. Zhang, X.-Z. Xing, A nomogram for predicting hospital mortality of critical ill patients with sepsis and cancer: A retrospective cohort study based on MIMIC-IV and eICU-CRD, BMJ Open 13 (2023) e072112. doi:10.1136/bmjopen-2023-072112.

[10] H. Tang, H. Hao, Y. Han, Personalized ICU mortality assessment by interpretable machine learning algorithms in patients with sepsis combined lung cancer: A population-based study and an external validation cohort, Frontiers in Oncology 15 (2025) 1661212. doi: 10.3389/fonc.2025.1661212.

[11] N. Hou, M. Li, L. He, B. Xie, L. Wang, R. Zhang, Y. Yu, X. Sun, Z. Pan, K. Wang, Predicting 30-days mortality for MIMIC-III patients with Sepsis-3: A machine learning approach using XGBoost, Journal of Translational Medicine 18 (2020) 462. doi:10.1186/s12967-020-02620-5.

[12] S. Li, R. Dou, X. Song, K. Y. Lui, J. Xu, Z. Guo, X. Hu, X. Guan, C. Cai, Developing an interpretable machine learning model to predict in-hospital mortality in sepsis patients: A retrospective temporal validation study, Journal of Clinical Medicine 12 (3) (2023) 915. doi:10.3390/jcm12030915.

[13] Y. Wang, Z. Gao, Y. Zhang, Z. Lu, F. Sun, Early sepsis mortality prediction model based on interpretable machine learning approach: Development and validation study, Internal and Emergency Medicine 20 (2025) 909–918. doi:10.1007/s11739-024-03732-2.

[14] C. Bao, F. Deng, S. Zhao, Machine-learning models for prediction of sepsis patients mortality, Medicina Intensiva 47 (6) (2023) 315–325. doi:10.1016/j.medin.2022.06.004.

[15] G. Zhang, F. Shao, W. Yuan, J. Wu, X. Qi, J. Gao, R. Shao, Z. Tang, T. Wang, Predicting sepsis in-hospital mortality with machine learning: A multi-center study using clinical and inflammatory biomarkers, European Journal of Medical Research 29 (1) (2024) 156. doi:10.1186/s40001-024-01756-0.

[16] E. Christodoulou, J. Ma, G. S. Collins, E. W. Steyerberg, J. Y. Verbakel, B. Van Calster, A systematic review shows no performance benefit of machine learning over logistic regression for clinical prediction models, Journal of Clinical Epidemiology 110 (2019) 12–22. doi:10.1016/j.jclinepi.2019.02.004.

[17] E. W. Steyerberg, A. J. Vickers, N. R. Cook, T. Gerds, M. Gonen, N. Obuchowski, M. J. Pencina, M. W. Kattan, Assessing the performance of prediction models: A framework for traditional and novel measures, Epidemiology 21 (1) (2010) 128–138. doi:10.1097/EDE.0b013e3181c30fb2.

[18] B. Van Calster, D. Nieboer, Y. Vergouwe, B. De Cock, M. J. Pencina, E. W. Steyerberg, A calibration hierarchy for risk models was defined: From utopia to empirical data, Journal of Clinical Epidemiology 74 (2016) 167–176. doi:10.1016/j.jclinepi.2015.12.005.

[19] T. P. A. Debray, Y. Vergouwe, H. Koffijberg, D. Nieboer, E. W. Steyerberg, K. G. M. Moons, A new framework to enhance the interpretation of external validation studies of clinical prediction models, Journal of Clinical Epidemiology 68 (3) (2015) 279–289. doi: 10.1016/j.jclinepi.2014.06.018.

[20] R. D. Riley, J. Ensor, K. I. E. Snell, T. P. A. Debray, D. G. Altman, K. G. M. Moons, G. S. Collins, External validation of clinical prediction models using big datasets from e-health records or IPD meta-analysis: Opportunities and challenges, BMJ 353 (2016) i3140. doi:10.1136/bmj.i3140.

[21] G. S. Collins, P. Dhiman, J. Ma, M. M. Schlussel, L. Archer, B. Van Calster, F. E. Harrell, G. P. Martin, K. G. M. Moons, M. van Smeden, M. Sperrin, G. S. Bullock, R. D. Riley, Evaluation of clinical prediction models (part 1): From development to external validation, BMJ 384 (2024) e074819. doi:10.1136/bmj-2023-074819.

[22] A. E. W. Johnson, L. Bulgarelli, L. Shen, A. Gayles, A. Shammout, S. Horng, T. J. Pollard, S. Hao, B. Moody, B. Gow, L.-W. H. Lehman, L. A. Celi, R. G. Mark, MIMIC-IV, a freely accessible electronic health record dataset, Scientific Data 10 (2023) 1. doi:10.1038/s41597-022-01899-x.

[23] T. J. Pollard, A. E. W. Johnson, J. D. Raffa, L. A. Celi, R. G. Mark, O. Badawi, The eICU collaborative research database, a freely available multi-center database for critical care research, Scientific Data 5 (2018) 180178. doi:10.1038/sdata.2018.178.

[24] S. M. Lundberg, S.-I. Lee, A unified approach to interpreting model predictions, in: Advances in Neural Information Processing Systems, Vol. 30, 2017, pp. 4765–4774. URL https://proceedings.neurips.cc/paper/2017/hash/8a20a8621978632d76c43dfd28b67767-Abstract.html

[25] G. S. Collins, K. G. M. Moons, P. Dhiman, R. D. Riley, A. L. Beam, B. Van Calster, M. Ghassemi, X. Liu, J. B. Reitsma, M. van Smeden, et al., TRIPOD+AI statement: Updated guidance for reporting clinical prediction models that use regression or machine learning methods, BMJ 385 (2024) e078378. doi:10.1136/bmj-2023-078378.

[26] K. G. M. Moons, J. A. A. Damen, T. Kaul, L. Hooft, C. Andaur Navarro, P. Dhiman, A. L. Beam, B. Van Calster, L. A. Celi, S. Denaxas, et al., PROBAST+AI: An updated quality, risk of bias, and applicability assessment tool for prediction models using regression or artificial intelligence methods, BMJ 388 (2025) e082505. doi:10.1136/bmj-2024-082505.

[27] F. Pedregosa, G. Varoquaux, A. Gramfort, V. Michel, B. Thirion, O. Grisel, M. Blondel, P. Prettenhofer, R. Weiss, V. Dubourg, J. Vanderplas, A. Passos, D. Cournapeau, M. Brucher, M. Perrot, E. Duchesnay, Scikit-learn: Machine learning in Python, Journal of Machine Learning Research 12 (2011) 2825–2830. URL https://jmlr.org/papers/v12/pedregosa11a.html

[28] M. Ghassemi, L. Oakden-Rayner, A. L. Beam, The false hope of current approaches to explainable artificial intelligence in health care, The Lancet Digital Health 3 (11) (2021) e745–e750. doi:10.1016/S2589-7500(21)00208-9.

