## Supplementary Material for "Interpretable machine learning prediction of in-hospital mortality in ICU patients with cancer and sepsis using first-day data: Development using MIMIC-IV and external validation in eICU-CRD"

### Table S1: Extended performance metrics for predictive models in the MIMIC-IV internal validation cohort.

| Model | AUROC (95% CI) | AUPRC | Threshold | Precision | Sensitivity | Specificity | F1 | Balanced accuracy | Accuracy | Brier |
|---|---|---|---|---|---|---|---|---|---|---|
| **GradientBoosting** | **0.8480 (0.8173–0.8755)** | **0.6984** | **0.2706** | **0.5567** | **0.7659** | **0.7689** | **0.6448** | **0.7674** | **0.7681** | **0.1346** |
| HistGradientBoosting | 0.8477 (0.8181–0.8759) | 0.6943 | 0.2524 | 0.5258 | 0.7951 | 0.7283 | 0.6330 | 0.7617 | 0.7466 | 0.1350 |
| Elastic-net logistic regression | 0.8389 (0.8070–0.8696) | 0.6795 | 0.2593 | 0.5329 | 0.7902 | 0.7375 | 0.6365 | 0.7639 | 0.7520 | 0.1386 |
| LASSO logistic regression | 0.8389 (0.8074–0.8684) | 0.6793 | 0.2562 | 0.5277 | 0.7902 | 0.7320 | 0.6328 | 0.7611 | 0.7480 | 0.1386 |
| Ridge logistic regression | 0.8361 (0.8023–0.8666) | 0.6750 | 0.2554 | 0.5060 | 0.8195 | 0.6969 | 0.6257 | 0.7582 | 0.7306 | 0.1400 |
| Balanced ridge logistic regression | 0.8348 (0.8009–0.8663) | 0.6725 | 0.4277 | 0.4970 | 0.8098 | 0.6895 | 0.6160 | 0.7496 | 0.7225 | 0.1664 |
| Balanced LASSO logistic regression | 0.8345 (0.8026–0.8647) | 0.6685 | 0.4702 | 0.5275 | 0.7951 | 0.7301 | 0.6342 | 0.7626 | 0.7480 | 0.1680 |
| AdaBoost | 0.8310 (0.7996–0.8596) | 0.6689 | 0.4386 | 0.4907 | 0.7707 | 0.6969 | 0.5996 | 0.7338 | 0.7172 | 0.1933 |
| Random forest | 0.8256 (0.7914–0.8559) | 0.6544 | 0.3483 | 0.4684 | 0.7951 | 0.6580 | 0.5895 | 0.7266 | 0.6957 | 0.1515 |
| Multilayer perceptron | 0.8215 (0.7862–0.8543) | 0.6526 | 0.0838 | 0.5331 | 0.7073 | 0.7652 | 0.6080 | 0.7363 | 0.7493 | 0.1753 |
| ExtraTrees | 0.8214 (0.7890–0.8527) | 0.6493 | 0.4210 | 0.5084 | 0.7366 | 0.7301 | 0.6016 | 0.7334 | 0.7319 | 0.1604 |
| *k*-nearest neighbors | 0.7702 (0.7294–0.8074) | 0.5760 | 0.1942 | 0.4700 | 0.6878 | 0.7061 | 0.5584 | 0.6970 | 0.7011 | 0.1721 |
| Decision tree | 0.7646 (0.7284–0.8021) | 0.5387 | 0.4292 | 0.4366 | 0.7561 | 0.6303 | 0.5536 | 0.6932 | 0.6649 | 0.2024 |
| Gaussian naive Bayes | 0.7341 (0.6945–0.7686) | 0.4595 | 1.0000 | 0.4918 | 0.5854 | 0.7708 | 0.5345 | 0.6781 | 0.7198 | 0.3525 |

Models are ordered by internal validation AUROC. Threshold-dependent metrics were calculated using model-specific operating thresholds selected from MIMIC-IV training-set out-of-fold predicted probabilities. The internal validation cohort was not used for threshold selection. GradientBoosting, shown in bold, was selected as the primary model according to the prespecified internal validation AUROC criterion. The most-frequent dummy classifier is not shown. AUPRC, area under the precision–recall curve; AUROC, area under the receiver operating characteristic curve; CI, confidence interval.

**Table S2: Extended performance metrics for predictive models under locked external validation in eICU-CRD.**

| Model | AUROC (95% CI) | AUPRC | Threshold | Precision | Sensitivity | Specificity | F1 | Balanced accuracy | Accuracy | Brier |
|---|---|---|---|---|---|---|---|---|---|---|
| Random forest | 0.7731 (0.7307–0.8123) | 0.6277 | 0.3483 | 0.4866 | 0.7713 | 0.6383 | 0.5967 | 0.7048 | 0.6792 | 0.1747 |
| ExtraTrees | 0.7600 (0.7130–0.8009) | 0.6003 | 0.4210 | 0.5221 | 0.6915 | 0.7187 | 0.5950 | 0.7051 | 0.7103 | 0.1865 |
| HistGradientBoosting | 0.7556 (0.7085–0.7969) | 0.6210 | 0.2524 | 0.4633 | 0.7394 | 0.6194 | 0.5697 | 0.6794 | 0.6563 | 0.1697 |
| **GradientBoosting** | **0.7483 (0.7026–0.7919)** | **0.6243** | **0.2706** | **0.4839** | **0.7181** | **0.6596** | **0.5782** | **0.6888** | **0.6776** | **0.1709** |
| Ridge logistic regression | 0.7473 (0.7006–0.7875) | 0.6057 | 0.2554 | 0.3781 | 0.8830 | 0.3546 | 0.5295 | 0.6188 | 0.5172 | 0.1931 |
| Balanced ridge logistic regression | 0.7433 (0.6973–0.7831) | 0.6038 | 0.4277 | 0.4330 | 0.8085 | 0.5296 | 0.5640 | 0.6690 | 0.6154 | 0.2246 |
| Balanced LASSO logistic regression | 0.7406 (0.6948–0.7816) | 0.6128 | 0.4702 | 0.4498 | 0.7394 | 0.5981 | 0.5594 | 0.6687 | 0.6416 | 0.2176 |
| Elastic-net logistic regression | 0.7389 (0.6898–0.7797) | 0.6016 | 0.2593 | 0.4774 | 0.6755 | 0.6714 | 0.5595 | 0.6735 | 0.6727 | 0.1775 |
| AdaBoost | 0.7378 (0.6910–0.7804) | 0.6035 | 0.4386 | 0.4846 | 0.6702 | 0.6832 | 0.5625 | 0.6767 | 0.6792 | 0.2106 |
| LASSO logistic regression | 0.7375 (0.6902–0.7779) | 0.6000 | 0.2562 | 0.4494 | 0.7553 | 0.5887 | 0.5635 | 0.6720 | 0.6399 | 0.1802 |
| $k$-nearest neighbors | 0.7001 (0.6498–0.7440) | 0.5547 | 0.1942 | 0.4332 | 0.6383 | 0.6288 | 0.5161 | 0.6336 | 0.6318 | 0.2030 |
| Decision tree | 0.6895 (0.6440–0.7360) | 0.4864 | 0.4292 | 0.4633 | 0.6383 | 0.6714 | 0.5369 | 0.6548 | 0.6612 | 0.2193 |
| Multilayer perceptron | 0.6649 (0.6165–0.7127) | 0.5266 | 0.0838 | 0.3392 | 0.8191 | 0.2908 | 0.4798 | 0.5550 | 0.4534 | 0.3994 |
| Gaussian naive Bayes | 0.5024 (0.5000–0.5060) | 0.3087 | 1.0000 | 0.3087 | 1.0000 | 0.0047 | 0.4718 | 0.5024 | 0.3110 | 0.6890 |

Models are ordered by external AUROC. All predictions used preprocessing pipelines, hyperparameters, and model-specific thresholds frozen after MIMIC-IV development. No feature reselection, model refitting, recalibration, or threshold re-estimation was performed using eICU-CRD. GradientBoosting, shown in bold, remained the prespecified primary model; random forest achieved the highest external AUROC as a secondary comparison. The most-frequent dummy classifier is not shown. AUPRC, area under the precision–recall curve; AUROC, area under the receiver operating characteristic curve; CI, confidence interval.

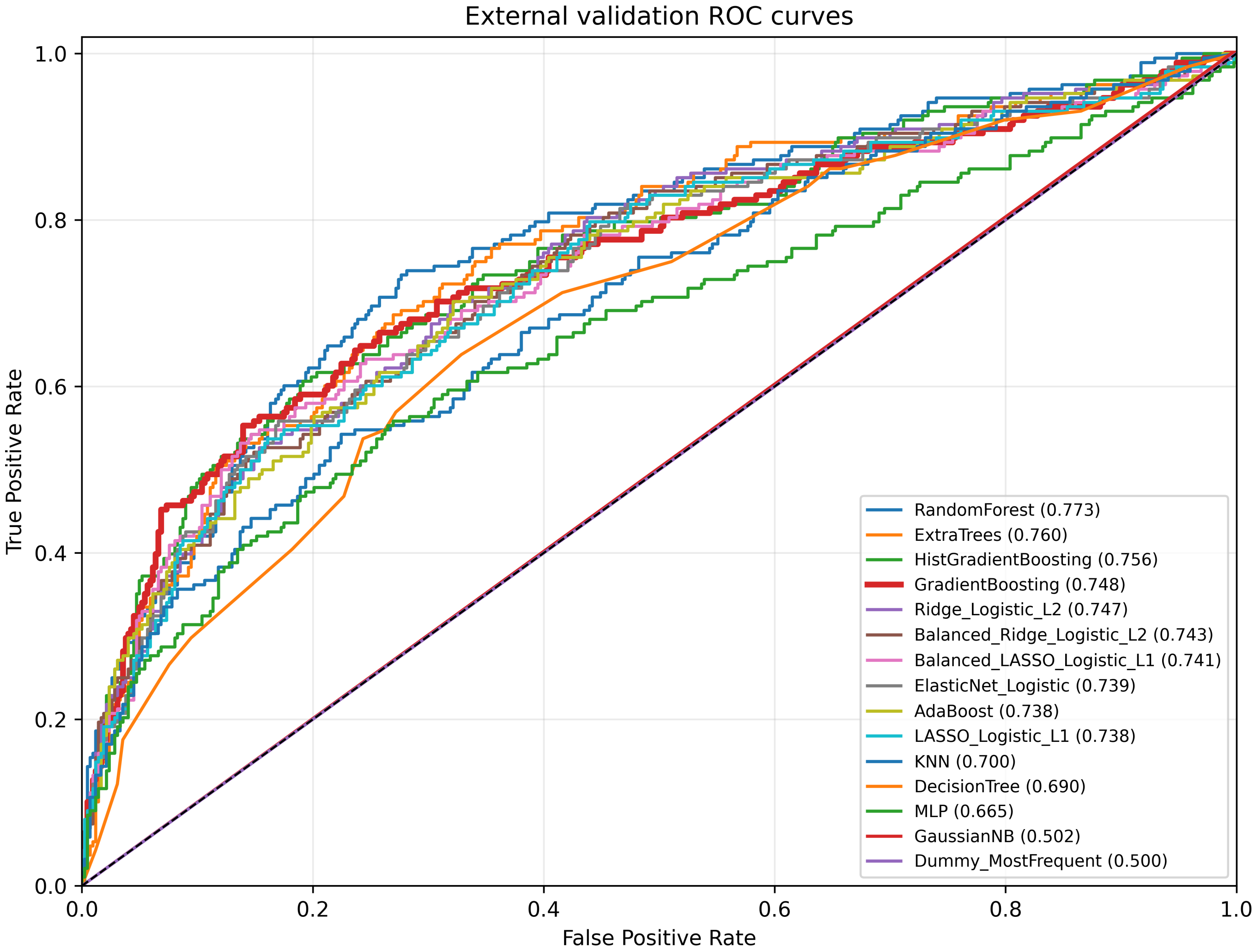


**Figure S1: Receiver operating characteristic curves for evaluated models and the dummy baseline under locked external validation in eICU-CRD.** All models were applied using preprocessing pipelines, fitted parameters, and feature definitions frozen after MIMIC-IV development, without external refitting or recalibration. AUROC, area under the receiver operating characteristic curve.

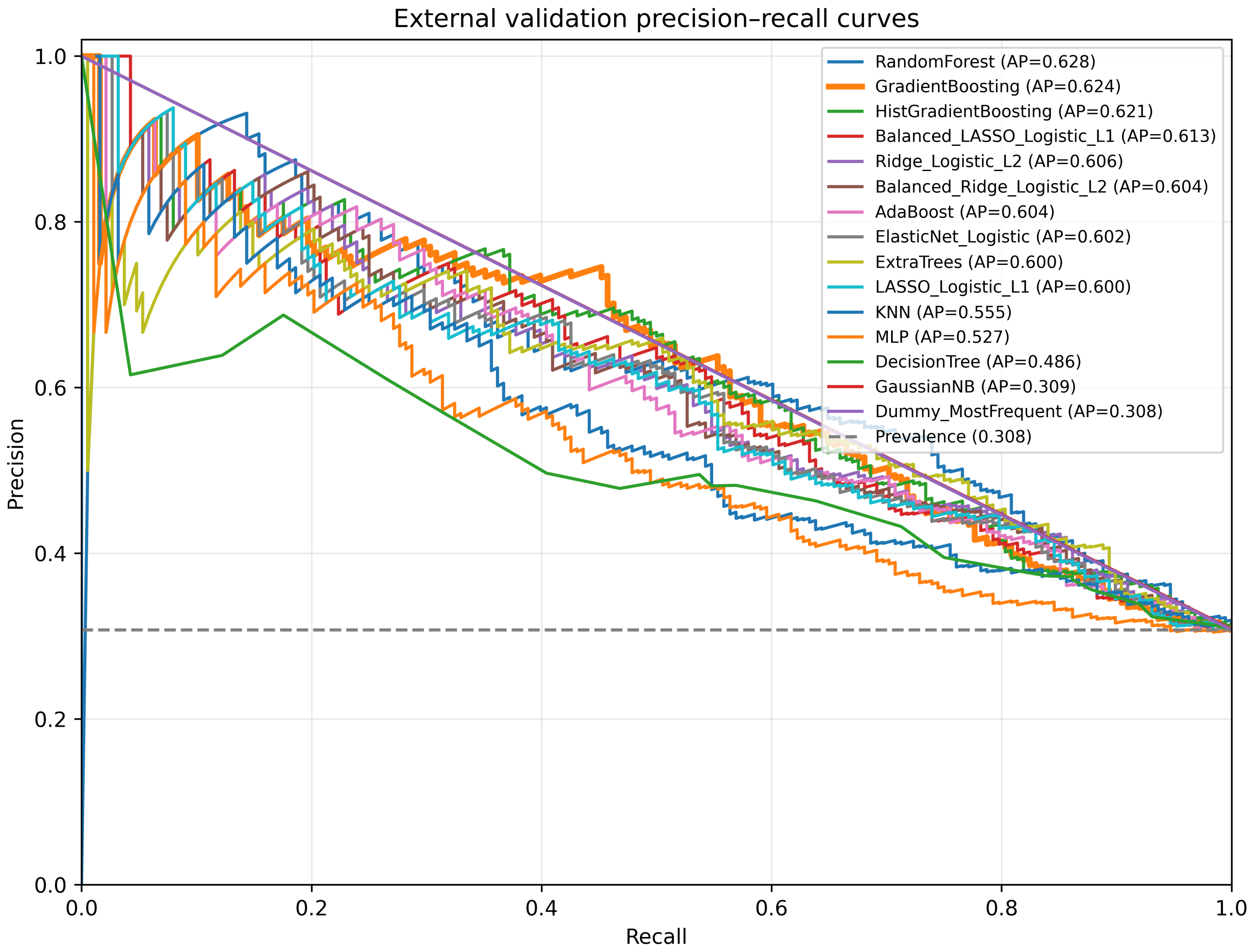


**Figure S2: Precision–recall curves for evaluated models and the dummy baseline under locked external validation in eICU-CRD.** All models were applied using preprocessing pipelines, fitted parameters, and feature definitions frozen after MIMIC-IV development, without external refitting or recalibration. AUPRC, area under the precision–recall curve.
